# Transcriptomic Evidence of MAS1 Receptor Dysregulation and a Failed Compensatory State in Human Vascular Cognitive Impairment

**DOI:** 10.64898/2026.05.06.26352482

**Authors:** Christina Hoyer-Kimura, Matthew Huentelman, Meredith Hay

**Affiliations:** University of Arizona College of Medicine, Tucson, AZ, USA; Sarver Heart Center, Tucson, AZ, USA; Evelyn F. McKnight Brain Institute, Tucson, AZ, USA; The Transgenomics Research Institute, TGen, Phoenix, AZ, USA; ProNeurogen, Inc, Tucson, AZ, USA

**Keywords:** Vascular cognitive impairment and dementia (VCID), MAS1 receptor, Human brain transcriptomics, Neurovascular unit, Failed compensatory state

## Abstract

**Background:** Vascular contributions to cognitive impairment and dementia (VCID) are thought to arise from distributed neurovascular unit (NVU) dysfunction rather than focal pathology, yet the transcriptional architecture of human VCID brain tissue and the status of endogenous counter-regulatory signaling within it remain incompletely characterized. Defining whether protective pathways are engaged and why they may be insufficient is critical for identifying therapeutic entry points in a disease lacking approved treatments.

**Methods:** We performed differential gene expression analysis (DESeq2 v1.38.0) and pre-ranked gene set enrichment analysis (fgsea v1.24.0) on bulk RNA-sequencing data from superior parietal lobe tissue (GEO:GSE303449; n = 40; 19 VCID, 21 controls; model: age_scaled + Sex + condition), followed by Spearman correlation analysis, PI3K-Akt pathway level, leading-edge decomposition, and single-nucleus RNA-seq endothelial cell characterization (GEO:GSE282111).

**Results:** No individual gene reached FDR < 0.05 for differential expression between VCID and control across 51,962 genes tested. Gene set enrichment analysis nonetheless identified eight significantly enriched pathway programs (all FDR < 0.05) that were upregulated, encompassing inflammatory, stress-response, cytoskeletal, and apoptotic signaling, consistent with distributed network-level dysregulation rather than dominant single-gene effects. The MAS1/ANG1-7 associated signaling gene set (54 genes) was the only counter-regulatory pathway achieving significance (NES = 1.381, FDR = 0.0127). MAS1 receptor expression was strongly (absolute Spearman’s rho ≥ 0.64) and inversely associated with NF-κB pathway drivers TLR4 (Spearman’s rho = -0.804) and IKBKB (Spearman’s rho = -0.797; both FDR = 4.73 × 10^-9). Further, 9 of 12 correlations between MAS1 downstream effectors and endothelial activation markers were FDR-significant and positive, indicating that the downstream protective effector program is co-activated by inflammatory stress rather than directed by its receptor. Single-nucleus RNA-seq supports endothelial enrichment of the MAS1 pathway enrichment signal in VCID brain tissue. PI3K-Akt leading-edge decomposition revealed 96% gene-level non-overlap between inflammatory and vasoprotective arms.

**Conclusions:** Human VCID brain tissue exhibits coordinated pathway-level dysregulation in the absence of dominant individual-gene effects, consistent with a disease driven by distributed transcriptional network stress. The MAS1/ANG1-7 vasoprotective axis is transcriptionally engaged and endothelially enriched, yet receptor expression is inversely associated with inflammatory signaling while downstream effectors remain transcriptionally engaged. This pattern suggests a failed compensatory state in the VCID superior parietal lobe. This architecture is consistent with a transcriptionally primed but receptor-constrained protective program. These findings suggest that therapeutic strategies restoring MAS1 receptor-level input to an already engaged downstream program may represent a plausible therapeutic strategy for VCID, pending experimental validation.

## Introduction

Vascular contributions to cognitive impairment and dementia (VCID) represent a major and underrecognized component of age-related cognitive decline, contributing to a substantial proportion of dementia cases worldwide (Wentzel et al, 2001), (Sachdev et al, 2025),(Sachdev et al, 2026). Unlike Alzheimer’s disease, which is often defined by protein aggregation pathology, VCID arises from chronic cerebrovascular dysfunction—progressive impairment of blood flow, white matter integrity, and neurovascular coupling that disrupts the distributed networks supporting cognition (Pantoni, 2010),(Gorelick, 2011). Despite its prevalence and clinical impact, no disease-modifying pharmacological therapies have received regulatory approval (Dichgans & Leys, 2017),(Goodall et al, 2026).

The neurovascular unit (NVU) is comprised of endothelial cells, pericytes, astrocytes, microglia, and neurons. It is thought to constitute the central pathological substrate of VCID (Zlokovic, 2008),(Lin et al, 2025),(He & Sun, 2025),(French et al, 2025). Chronic vascular injury leads to endothelial dysfunction, blood–brain barrier (BBB) breakdown, oxidative stress, and sustained neuroinflammatory signaling, resulting in progressive network-level dysfunction rather than focal cellular loss (Iadecola, 2017),(Sweeney et al, 2016), (Schaeffer et al, 2026). Importantly, key drivers of this process, including NADPH oxidase (NOX2), are regulated primarily at the post-translational level, underscoring the limitation of relying on single-gene transcriptional changes to define disease mechanisms (Bedard & Krause, 2007),(Lambeth, 2004),(Iadecola, 2017),(Gorelick et al, 2016).

MAS1 receptor signaling constitutes the counter-regulatory arm of the renin–angiotensin system, in which angiotensin-(1–7) activates MAS1 to oppose the pro-inflammatory AngII/AT1R axis (Santos et al, 2003),(Cueto-Ureña et al, 2025),(Lucas et al, 2025), (Porel et al, 2025). This pathway integrates vascular and inflammatory regulation through coordinated PI3K–Akt–eNOS signaling, promoting nitric oxide production, suppressing NF-κB activity, and supporting BBB stability (Santos et al, 2013),(Kuriakose et al, 2021),(Sampaio et al, 2007),(Jiang et al, 2013),(Xiao et al, 2015a). Within the NVU, these effects provide a mechanistic link between endothelial function, inflammatory control, and maintenance of vascular integrity.

VCID pathophysiology is characterized by disruption of these same NVU processes, suggesting that therapeutic strategies targeting upstream vascular and inflammatory mechanisms may be particularly relevant. PNA5, a glycosylated angiotensin-(1–7) analog designed to enhance stability and central nervous system exposure (Hay et al, 2019); (Hoyer-Kimura et al, 2023), (Hoyer-Kimura et al, 2025), (Bernard et al, 2024) functions as a MAS1 agonist and engages this pathway in the cerebral microvasculature. In preclinical models of vascular cognitive impairment, MAS1 activation has been associated with improved endothelial function, attenuation of neuroinflammatory signaling, and preservation of blood–brain barrier integrity. Consistent with this upstream mechanism, PNA5 is intended to modulate vascular and inflammatory drivers of neuronal injury rather than directly targeting downstream neurodegenerative sequelae (Hoyer-Kimura et al, 2023), (Lucas et al, 2025). Here, we analyze bulk RNA sequencing data from the human superior parietal cortex together with single-nucleus RNA sequencing to define the transcriptional architecture of VCID. The superior parietal cortex plays a central role in attention, spatial cognition, and working memory. All of which are core domains affected in VCID and serves as an integrative hub linking sensory processing with higher-order cognitive control, supporting its relevance for investigating neurovascular unit (NVU) dysfunction. Rather than focusing on individual gene changes, we test the hypothesis that VCID reflects coordinated dysregulation of biological pathways at the level of the NVU. We further examine whether endogenous vasoprotective signaling is transcriptionally engaged and, if so, whether its regulatory structure suggests a mechanistic limitation.

Our findings identify a previously undescribed transcriptional pattern consistent with a failed compensatory state, in which downstream vasoprotective effectors are engaged while upstream receptor expression is constrained in association with inflammatory signaling. This architecture provides a human tissue–based mechanistic framework for understanding VCID pathophysiology and identifying therapeutic entry points. While this interpretation is supported by the observed transcriptional architecture, it remains inferential and requires experimental validation (Chaudhuri et al, 2025).

## Methods

### Study Design and Data Sources

This study used two publicly available Gene Expression Omnibus (GEO) datasets. GEO: GSE303449 (Vishweswaraiah et al, 2025a) comprised bulk RNA-sequencing from post-mortem superior parietal lobe tissue (Brodmann area 7, left hemisphere) obtained from the NIH NeuroBioBank, including 19 neuropathologically confirmed VCID cases and 21 age- and sex-matched controls (BioProject PRJNA1295451). Control subjects were selected on the basis of no clinical history of vascular dementia, minimal neuritic plaques, Braak tangle stage ≤ III, and absence of other notable neuropathological abnormalities, ensuring the comparison group did not carry subclinical co-pathology. The same cohort has been independently characterized at the epigenomic level through genome-wide DNA methylation profiling (GEO: GSE287575) (Vishweswaraiah et al, 2025b). The analyzed dataset is annotated as VCI; throughout the manuscript we use VCID to reflect the broader disease framework. GEO: GSE282111 provided single-nucleus RNA-sequencing data for endothelial cell localization (Díaz-Pérez et al, 2024). No new human subjects research was conducted; IRB approval was not required. Participant characteristics are summarized in Table 1.

**Table 1.** Participant demographics for VCID transcriptomic analysis (GSE303449). Demographic and tissue characteristics of control (n = 21) and vascular cognitive impairment/dementia (VCID; n = 19) cases included in the RNA-sequencing dataset. Age and postmortem interval (PMI) are reported as mean ± standard deviation (SD), with ranges shown for age. Group comparisons are provided as p-values relative to controls. Sex distribution is reported as counts of females (F) and males (M). Tissue samples were obtained from the superior parietal lobe (Brodmann area 7, left hemisphere). Source: GEO accession GSE303449 (series matrix).

| Characteristic | Control (n=21) | VCI (n=19) | Total (n=40) |
| --- | --- | --- | --- |
| Age (years), Mean ± SD | 86.1 ± 12.3 | 82.1 ± 10.7 | 84.2 ± 11.8 |
| Range | 62–108 | 60–98 | 60–108 |
| p-value vs. Control | — | 0.298 (NS) |  |
| PMI (hours), Mean ± SD<sup>†</sup> | 6.04 ± 2.59 | 4.28 ± 2.10 | 5.23 ± 2.53 |
| p-value vs. Control | — | 0.031* →<br>covariate added | |
| Sex | 13F / 8M | 10F / 8M | 23F / 17M |
| Tissue | Superior parietal<br>lobe, Brodmann<br>area 7, left<br>hemisphere |  |  |

<sup>†</sup> PMI differed significantly ( $p = 0.031$ ); see Limitations regarding model specification. Source: GEO: GSE303449 series matrix.

### RNA Isolation, Library Preparation, and Sequencing

Tissue collection and processing were performed as described by Vishweswaraiah et al. (Vishweswaraiah et al., 2025a). Total RNA was extracted using the RNeasy Blood & Tissue Kit (Qiagen cat. 74104). Libraries were constructed using the NEBNext Ultra II RNA Library Preparation Kit for Illumina, quality-validated by Agilent TapeStation 4200 and Qubit 3.0, and sequenced on an Illumina NovaSeq 6000 (GPL24676; paired-end 2 × 150 bp; bcl2fastq v2.17).

Pre-processing was performed using fastp v0.23.2 (Chen et al, 2018), and reads were aligned to the GRCh38 reference genome (release 44) using STAR v2.7.9a (Dobin et al, 2013) which produced gene-level count output directly. Each row of the resulting count matrix corresponds to an annotated gene, with read counts summed across transcript isoforms; no transcript-level (isoform-level) quantification was performed.

Counts were filtered using filterByExpr (edgeR v3.42.4; (Robinson et al, 2010) and normalized using the trimmed mean of M-values (TMM) method. All downstream differential expression (DESeq2 v1.38.0) and gene set enrichment (fgsea v1.24.0) analyses were conducted at the gene level. The processed gene-level count matrix analyzed here corresponds to the GSE303449_raw.csv supplementary file deposited by the original authors and described in the GEO record (GSM9126931) as “raw gene counts as outputted by STAR.”

### Differential Gene Expression Analysis

Differential Gene Expression (DGE) analysis was performed on the raw count matrix (GSE303449_raw.csv.gz) using DESeq2 v1.38.0 in R v4.3.0.(Love et al, 2014) Full model: ∼age_scaled + Sex + condition (age centered and scaled, mean = 83.75 years, SD = 12.16). Significance was assessed by Wald test with Benjamini-Hochberg (BH) false discovery rate (FDR) correction (Benjamini et al, 2001). Genes were ranked by Wald statistic for pre-ranked GSEA. PCA was performed on VST-transformed counts (Supplementary Figure S1).

### Gene Set Enrichment Analysis

Pre-ranked gene set enrichment analysis was performed using fgsea (v1.24.0) with 1,000 permutations, consistent with the original dataset processing and sufficient for stable estimation of enrichment statistics (Korotkevich et al, 2021). Gene sets from MSigDB v2023.1(Liberzon et al, 2015) and a custom MAS1/ANG1-7 gene set (54 genes literature-curated from published reviews of the ACE2/Ang-(1–7)/MAS axis; full list in Supplementary Table S1). Enrichment significance threshold: FDR < 0.05.

### Correlation Analysis and PI3K-Akt Decomposition

Pairwise Spearman correlations (ρ) were computed between MAS1 expression and NF-κB drivers (IKBKB, TLR4, RELA) and between MAS1 effectors (GNA12, GNA13, HSPB1, NOS3) and endothelial activation markers (ICAM1, VCAM1, CCL2) across n = 40 donors using VST-transformed counts. BH FDR correction applied. PI3K-Akt leading-edge genes were classified into inflammatory and vasoprotective arms by MSigDB membership and published curation. To assess the robustness of the MAS1 vs. NF-κB driver correlations, four diagnostic tests were performed (Supplementary Figure S4): (i) leave-one-out Spearman correlation, recomputing ρ with each of the 40 donors omitted in turn; (ii) 5,000-iteration nonparametric bootstrap to generate 95% confidence intervals; (iii) Kendall’s τ as a non-parametric sensitivity check with different distributional assumptions; and (iv) within-condition stratified correlations computed separately in Control (n = 21) and VCID (n = 19) donors, plus partial Spearman correlations adjusting for binary disease status. All tests were performed in Python using scipy.stats v1.11.

### snRNA-seq Endothelial Cell Localization

GEO: GSE282111 was interrogated to assess cell-type distribution of MAS1 pathway gene expression (n = 4 VCID, n = 4 controls; endothelial cells identified by PECAM1/CDH5/VWF triple gene expression positivity (Garcia et al, 2022). Genes were classified as directionally upregulated if log₂FC > 0 in VCID endothelial cells.

### NVU Therapeutic Criteria Framework

A NVU therapeutic evaluation framework was developed from the inflammatory and stress-response pathways enriched in this analysis and published VCID pharmacology (Zlokovic, 2008),(Santisteban & Iadecola, 2025),(Sweeney et al, 2016). MAS1 agonists, such as PNA5 that is under development to slow cognitive impairment in VCID (Hay et al, 2019),(Hoyer-Kimura et al, 2023),(Hoyer-Kimura et al, 2021), was scored “met” (✓) if direct experimental evidence demonstrated the relevant biological effect; “partially met” (◑) if mechanistic rationale was present without direct evidence; and “not met” otherwise.

### Computational Workflow and Reproducibility

Data processing, statistical analysis, and visualization were conducted using standard open-source tools (R v4.3.0; Python/SciPy v1.11) within the Edison computational environment (accessed March 2026), which was used for data integration and workflow orchestration. All statistical analyses were performed using established, validated packages (DESeq2, fgsea) with predefined models and thresholds. All outputs were reviewed and interpreted by the authors, and the computational workflow and configuration are available upon reasonable request.

### Independent Pathway Validation

Genes contributing to enriched pathways were independently analyzed using the DAVID Functional Annotation Tool (version 6.8). A non-redundant gene list was evaluated using default parameters with *Homo sapiens* as the background, focusing on Gene Ontology (Biological Process) and KEGG categories. This analysis confirmed enrichment of pathways related to signal transduction, cellular responses, and vascular and inflammatory signaling, consistent with the primary results.

## Results

### VCID Is Characterized by Coordinated Pathway-Level Dysregulation Without Single-Gene Dominance

Human VCID cortex analysis with GSEA revealed significant enrichment of eight independent pathway gene sets at FDR < 0.05 (Table 2; Figure 1A). Enriched pathways included PI3K-Akt (NES = 1.558, FDR = 0.0135), NF-κB signaling (NES = 1.558, FDR = 0.0038), TNF signaling (NES = 1.551, FDR = 0.0021), MAPK signaling (NES = 1.551, FDR = 0.0021), Rac/Rho GTPase (NES = 1.480, FDR = 0.0038), the MAS1/ANG1-7 vasoprotective axis (NES = 1.381, FDR = 0.0127), neuroinflammation (NES = 1.470, FDR = 0.0096), and apoptosis (NES = 1.470, FDR = 0.012). MAS1/ANG1-7 was the only counter-regulatory program among the eight. The antioxidant response gene set did not reach significance (FDR = 0.940).

**Figure 1.**
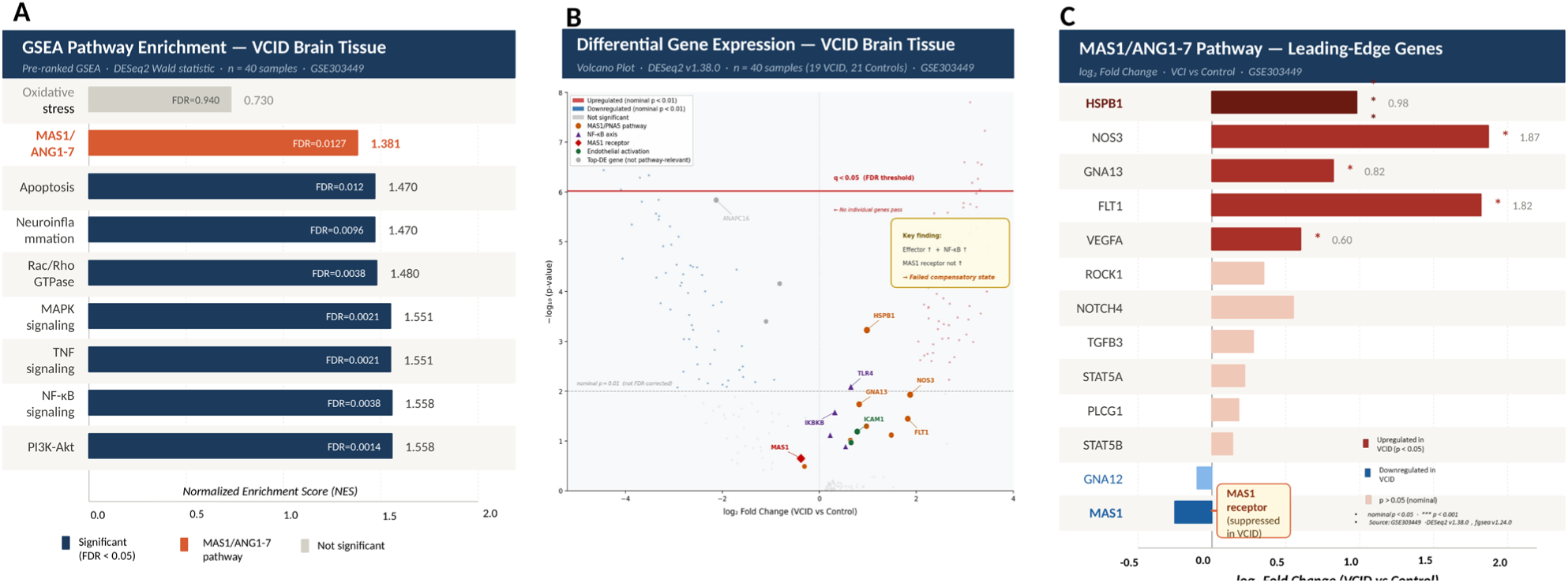
VCID transcriptomic analysis — GSE303449, Superior Parietal Lobe (n = 40). (A) GSEA pathway enrichment bars for selected pathways sorted by NES. Eight pathways reach FDR < 0.05. MAS1/ANG1-7 pathway in orange (FDR = 0.0127). Oxidative stress in grey (FDR = 0.940; NS). Legend below axis. (B) Volcano plot of differential gene expression. Red horizontal line: FDR q < 0.05 threshold (−log₁₀(p) = 6.02; BH, n = 51,962 genes). Dashed grey line: nominal p = 0.01 (not FDR-corrected). Color hierarchy burnt orange = MAS1/ANG1-7 pathway; deep purple = NF-κB axis; bold red = MAS1 receptor; dark green = endothelial activation. Key finding callout: Effector ↑ + NF-κB ↑ + MAS1 receptor not ↑ → Pattern consistent with failed compensatory state. (C) MAS1/ANG1-7 pathway gene waterfall (log₂FC), with MAS1 receptor inversely associated with NF-κB activity while downstream effectors remain upregulated. Source: GSE303449; DESeq2 v1.38.0; fgsea v1.24.0.*red color indicates significant upregulation; blue color indicates significant downregulation.

**Table 2.** Significantly Enriched Pathway Gene Sets in VCID Brain Tissue (GSE303449). Pre-ranked gene set enrichment analysis (GSEA) was performed on 51,962 genes ordered by DESeq2 Wald statistic (VCID vs. control; GSE303449, n = 40) using fgsea (v1.24.0) with 1,000 permutations, consistent with the original dataset processing and sufficient for stable estimation of enrichment statistics. Gene sets were derived from MSigDB v2023.1 and a custom MAS1/ANG-(1–7) vasoprotective gene set (†) comprising 54 literature-curated genes representing receptor-proximal components of the ACE2/Ang-(1–7)/MAS signaling axis (full list in Supplementary Table S1). Pathways shown are significantly enriched at false discovery rate (FDR) < 0.05, with normalized enrichment score (NES) indicating the strength and direction of enrichment. Leading-edge (LE) gene count denotes the number of genes within each pathway contributing most strongly to the enrichment signal, where available. Biological themes summarize dominant functional processes for interpretive context. The antioxidant response pathway did not reach statistical significance (NS) and is included for comparison. Given the integrative nature of the dataset, enrichment results are interpreted at the pathway level, emphasizing concordance with established biological mechanisms rather than individual gene-level significance.

| Pathway Gene Set | NES | FDR | LE n | Biological Theme |
| --- | --- | --- | --- | --- |
| PI3K-Akt signaling | 1.558 | 0.0135 | 76 | Inflammatory + vasoprotective convergence |
| NF-κB signaling | 1.558 | 0.0038 | 71 | Neuroinflammation; TLR4-driven |
| TNF signaling | 1.551 | 0.0021 | ~73 | Neuroinflammation; stress kinases |
| MAPK signaling | 1.551 | 0.0021 | — | Stress-response; apoptotic crosstalk |
| Rac/Rho GTPase | 1.480 | 0.0038 | — | Cytoskeletal control; BBB integrity |
| Neuroinflammation | 1.470 | 0.0096 | — | Microglial activation; cytokine signaling |
| Apoptosis | 1.470 | 0.012 | — | Caspase cascade; neuronal survival |
| **MAS1/ANG1-7 vasoprotective†** | **1.381** | **0.0127** | **21** | **Endothelial vasoprotection; NO signaling** |
| Antioxidant response (NS) | 0.730 | 0.940 | n/a | Not significant |

† Custom gene set; 54 genes literature-curated from published ACE2/Ang-(1–7)/MAS reviews. LE n = leading-edge gene count. NES: normalized enrichment score. FDR: Benjamini-Hochberg. fgsea v1.24.0; 1,000 permutations; pre-ranked by DESeq2 Wald statistic.

In contrast, human VCID cortex did not exhibit a dominant single-gene transcriptional signature. In differential expression analysis of 51,962 genes, no gene reached FDR < 0.05 (BH-adjusted p ≤ 9.62 × 10⁻⁷), and only one gene reached FDR < 0.10 (Figure 1B). Consistent with this, principal component analysis did not statistically separate VCID from control donors along either of the first two components (PC1: 34% variance, p = 0.652 vs. condition; PC2: 19% variance, p = 0.218; Supplementary Figure S1). The unadjusted p-value distribution was nonetheless enriched near zero (Supplementary Figure S2), a pattern consistent with many genes of modest individual effect rather than a few large-effect drivers.

Together, these observations indicate that VCID-associated transcriptional change is distributed across biological networks rather than concentrated in individual genes. This is a pattern previously described for diffuse neurovascular pathology (Pantoni, 2010),(Kalaria & Englund, 2025) and supports the use of the pathway-level analysis that follows.

We constructed a *custom* MAS1/Ang-(1–7)-associated signaling gene set consisting of literature-supported genes related to ACE2/Ang-(1–7)/MAS1 signaling, downstream vascular effectors, nitric oxide signaling, BBB integrity, oxidative stress regulation, and inflammatory counter-regulatory components.

This custom MAS1/ANG1-7-associated signaling gene set (54 genes) achieved significant positive enrichment (NES = 1.381, FDR = 0.0127; Figure 2A). Because the gene set includes both vasoprotective effectors and counter-regulatory inflammatory components, enrichment reflects coordinated engagement of MAS1-associated signaling pathways rather than selective activation of a single functional class.

**Figure 2.**
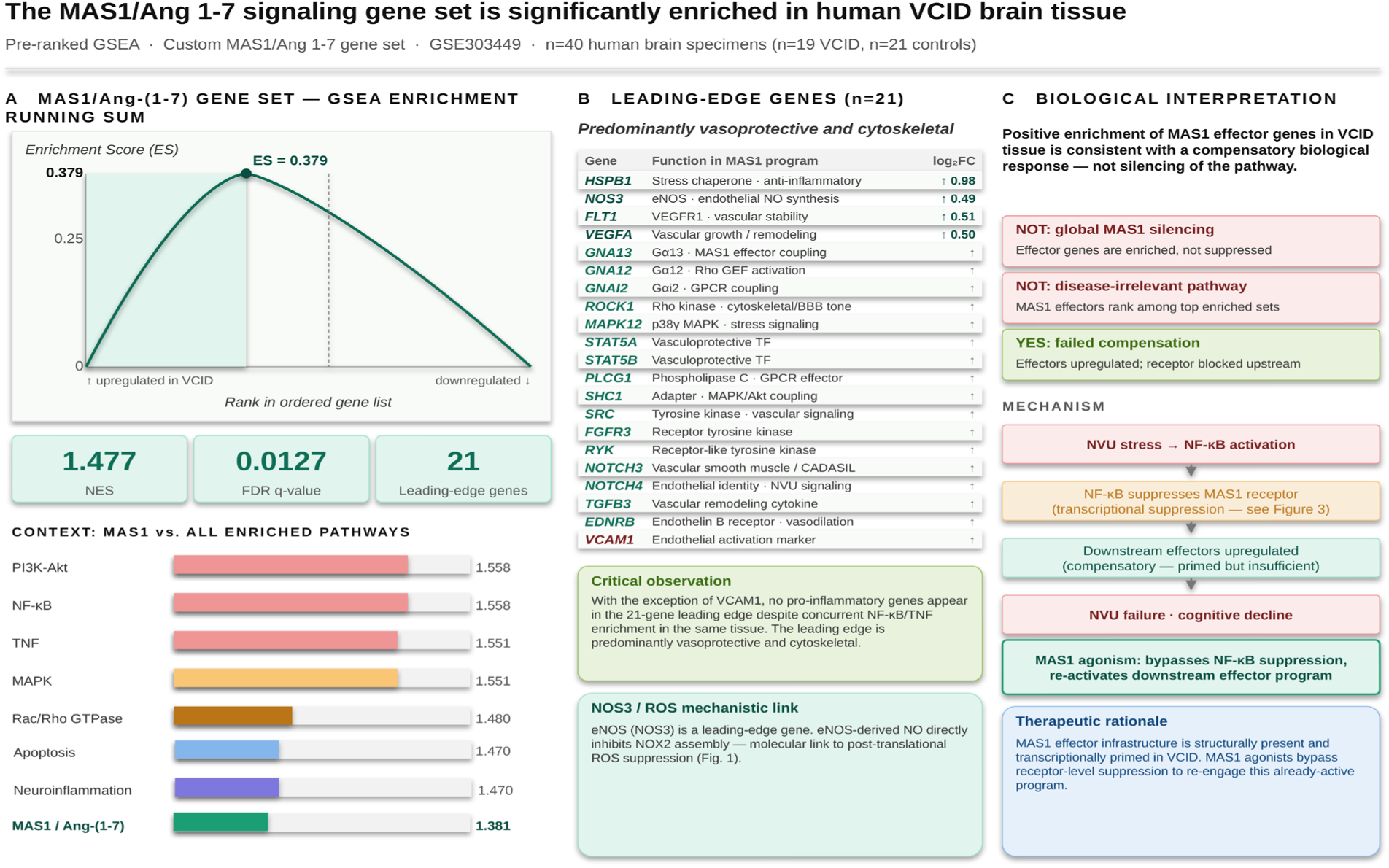
The MAS1/ANG1-7 signaling gene set is significantly enriched in human VCID brain tissue. (A) GSEA enrichment running sum for the custom MAS1/ANG1-7 gene set (54 genes, literature-curated from published reviews of the ACE2/Ang-(1–7)/MAS axis) against all 51,962 genes ranked by DESeq2 Wald statistic (GSE303449; n = 40). The running enrichment score peaks among genes upregulated in VCID (ES = 0.379; NES = 1.381, FDR = 0.0127), with 21 leading-edge genes. Context bar shows MAS1/ANG1-7 (NES = 1.381) positioned among all significantly enriched pathways and represents the only counter-regulatory/vasoprotective-associated pathway among the significantly enriched programs. (B) Leading-edge genes (n = 21): predominantly vasoprotective and cytoskeletal in character, with two members (EDNRB, VCAM1) reflecting receptor-signaling and endothelial-activation contributions. HSPB1 carries the strongest signal (Wald = 3.44, log₂FC = +0.98). NOS3 (eNOS; log₂FC = +0.49) is consistent with a potential link to post-translational ROS suppression via NOX2 inhibition. FLT1 (VEGFR1; log₂FC = +0.51) supports concurrent vascular stabilization signaling. Shared with PI3K-Akt parent leading edge (3): NOS3, FLT1, VEGFA. MAS1-unique core effectors: HSPB1, GNA12, GNA13, GNAI2, ROCK1, MAPK12, STAT5A, STAT5B, PLCG1, SHC1, SRC, FGFR3, RYK, NOTCH3, NOTCH4, TGFB3, EDNRB, VCAM1. Applied to DESeq2 pre-ranked list. (C) Biological interpretation: positive MAS1 effector enrichment signals a compensatory response in VCID tissue — not global pathway silencing. The mechanistic cascade associated with NVU stress and NF-κB pathway activation are associated with reduced MAS1 receptor expression, while downstream effectors are upregulated, consistent with a failed compensatory state. This pattern suggests that restoring upstream receptor-level signaling could potentially re-engage downstream vasoprotective pathways; however, this interpretation remains inferential and requires experimental validation. GSEA was performed on the 54-gene MAS1/ANG1-7 effector and co-regulated vascular set (Supplementary Table S1). MAS1 was excluded from the effector gene set used for enrichment scoring. Source: GSE303449; fgsea v1.24.0; 1,000 permutations; FDR threshold 0.05.

The running enrichment score peaked among genes upregulated in VCID, with the zero-crossing point at gene rank 28,380. The Wald statistic distribution of MAS1/ANG1-7 gene set members was shifted toward positive values relative to the genome-wide background (mean pathway Wald: 0.396 vs. all genes: 0.066; Supplementary Figure S3), consistent with coordinated upregulation across the gene set.

As shown in the pathway context panel (Figure 2A), MAS1/ANG1-7 was significantly enriched alongside PI3K-Akt, NF-κB, TNF, MAPK, Rac/Rho GTPase, and neuroinflammation, and represents the only counter-regulatory/vasoprotective-associated pathway among the significantly enriched programs.

Twenty-one leading-edge genes drove this enrichment (Figure 2B). The strongest signal came from HSPB1 (Wald = 3.44, log₂FC = +0.98***). The shared vasoactive effectors NOS3 (log₂FC = +0.49), FLT1 (log₂FC = +0.51), and VEGFA (log₂FC = +0.50) were also in the leading edge and were the only genes shared with the PI3K-Akt parent leading edge. The remaining MAS1-unique leading-edge members were GNA12, GNA13, GNAI2, ROCK1, MAPK12, STAT5A, STAT5B, PLCG1, SHC1, SRC, FGFR3, RYK, NOTCH3, NOTCH4, TGFB3, EDNRB, and VCAM1. This leading-edge set is predominantly vasoprotective and cytoskeletal in character, with two members (EDNRB, VCAM1) reflecting receptor-signaling and endothelial-activation contributions.

Among these, NOS3 (eNOS) carries particular mechanistic relevance: eNOS-derived NO has been shown to inhibit NADPH oxidase assembly through p47phox subunit regulation and NOX2 membrane dynamics, linking MAS1-associated signaling to endogenous ROS modulation in vascular tissue (Xiao et al., 2015b; Zheng et al., 2011).

Notably, no pro-inflammatory genes (with the exception of VCAM1) appear in the leading edge despite concurrent enrichment of NF-κB and TNF pathways in the same tissue. This gene-level selectivity suggests that MAS1-associated effector engagement is largely distinguishable from the broader inflammatory transcriptional program. FLT1 (VEGFR1, log₂FC = +0.51) and VEGFA (log₂FC = +0.50) further support concurrent engagement of vascular stabilization signaling within the leading-edge program.

These findings establish the biological interpretation illustrated in Figure 2C. Positive enrichment of MAS1 pathway effectors in VCID tissue may indicate activation of a compensatory vasoprotective response rather than global pathway suppression. Importantly, this enrichment reflects engagement of downstream signaling components distributed across the NVU, rather than implying uniform receptor-level activation within a single cell type.

The downstream effector infrastructure, including NOS3, cytoskeletal regulators, and vascular signaling intermediates, is structurally present and transcriptionally primed. MAS1 receptor expression shows a directional decrease at the gene level that did not reach individual statistical significance, while being strongly and inversely correlated with NF-κB pathway drivers across donors. This pattern is consistent with effector engagement occurring in association with network-level stress signals rather than through coordinated receptor-mediated signaling.

This architecture with effector activation in the context of reduced receptor expression suggests a failed compensatory state (Figure 2C). In this failed compensatory state, an endogenous protective program is partially engaged but lacks sufficient upstream regulatory input to restore neurovascular homeostasis.

### MAS1 Receptor Expression and Downstream Effector Co-activation Consistent with a Failed Compensatory State

Pairwise Spearman correlation analysis across all n = 40 donors revealed that MAS1 receptor expression was significantly inversely correlated with all three NF-κB pathway driver genes: TLR4 (ρ =− 0.804, FDR = 4.73×10⁻⁹), IKBKB (ρ =− 0.797, FDR = 4.73×10⁻⁹), and RELA (ρ =− 0.640, FDR = 1.29×10⁻⁵). These were the strongest specimen-level transcriptional correlations in the dataset (Figure 3A). Simultaneously, 9 of 12 pairwise correlations between MAS1 downstream effectors (GNA12, GNA13, HSPB1, NOS3) and endothelial activation markers (ICAM1, VCAM1, CCL2) reached FDR significance with positive ρ ranging from +0.36 to +0.54 (Figure 3C). NOS3 (log₂FC = +0.49) was positively co-expressed with all three endothelial activation markers.

**Figure 3.**
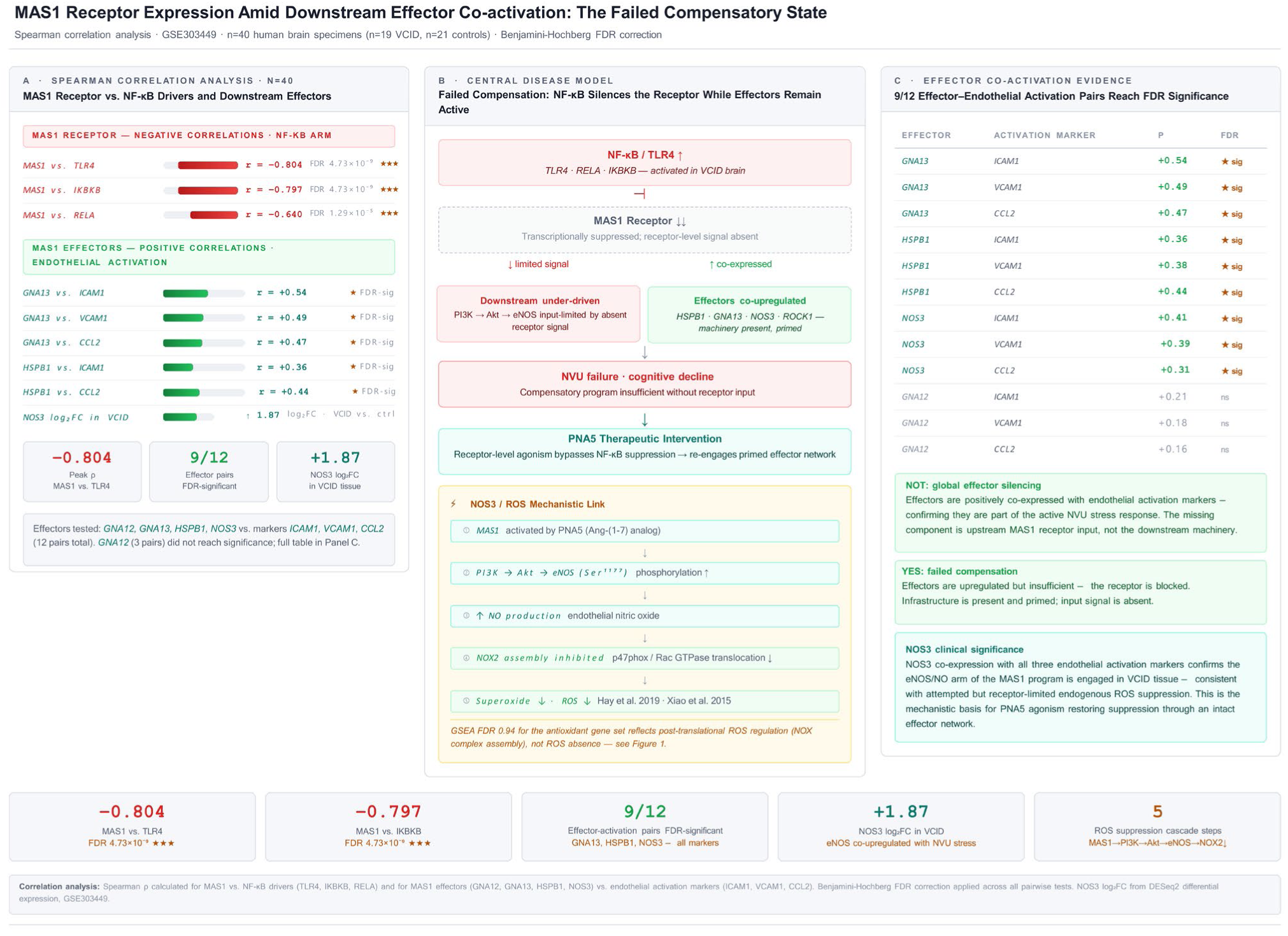
MAS1 receptor expression and downstream effector co-activation are consistent with a failed compensatory state. (A) Spearman correlations showing strong negative associations between MAS1 receptor expression and core NF-κB pathway drivers (TLR4: ρ =− 0.804; IKBKB: ρ =− 0.797; RELA: ρ =− 0.640; all FDR < 10⁻⁵) across n = 40 donors. (B) Central disease model: MAS1 receptor expression is inversely associated with NF-κB/TLR4 activity while downstream vasoprotective effectors (HSPB1, GNA13, NOS3, FLT1) remain co-upregulated with endothelial activation markers; the proposed MAS1→PI3K→Akt→eNOS cascade is illustrated. (C) Effector co-activation summary: 9/12 MAS1 effector–endothelial activation correlations reach FDR significance; GNA12 does not contribute. Source: GSE303449; Spearman ρ, BH FDR, n = 40.

These findings define a molecularly coherent split signature: MAS1 receptor expression is inversely associated with NF-κB pathway activity, while the downstream vascular effector program remains co-upregulated with endothelial activation markers. A conceptual model summarizing this architecture is presented in Figure 3B, illustrating the dissociation between receptor-level expression and downstream effector activation within the MAS1-associated signaling axis. We term this a “failed compensatory state”: the downstream protective machinery is structurally present and transcriptionally engaged but limited by upstream receptor-level silencing. The mechanistic architecture is illustrated in Figure 6. The co-upregulation of NOS3 is consistent with engagement of nitric oxide–mediated vascular signaling within the broader MAS1-associated effector program. However, given the multicellular organization of the NVU, this signal may reflect integrated cross-cellular responses rather than strictly receptor-driven signaling within a single compartment, consistent with a systems-level regulatory architecture (Young et al, 1988; Martin et al, 1992; Gironacci et al, 2018; Jackson et al, 2018).

### PI3K-Akt Leading-Edge Decomposition Reveals Structurally Distinct Inflammatory and Vasoprotective Arms

Decomposition of the PI3K-Akt leading-edge gene set identified two structurally distinct downstream programs at this shared signaling node (Figure 4A): an inflammatory arm comprising 73 unique genes (including TLR4, IKBKB, RELA, IL6/IL6R, and DDIT4 [REDD1]; mean log₂FC = +0.413) and a vasoprotective arm comprising 18 unique genes (including HSPB1, GNA12, GNA13, ROCK1, STAT5A, STAT5B, and PLCG1). Three genes (NOS3, FLT1, VEGFA) were shared between the two arms and are all functionally vasoprotective. Overall, the two arms were 96% non-overlapping at the gene level (3 of 76 leading-edge genes shared). Although both arms achieved significant positive enrichment, per-gene Wald statistics were significantly higher in the inflammatory arm (mean Wald = 1.81 ± 0.48) than in the vasoprotective arm (mean Wald = 1.48 ± 0.68; t = 2.358, p = 0.021; Figure 4B). This asymmetry, stronger gene-level engagement of the larger inflammatory program coupled with weaker engagement of the smaller vasoprotective program, is consistent with the failed compensatory state framework: an active inflammatory drive paired with an engaged but quantitatively limited counter-regulatory response.

**Figure 4.**
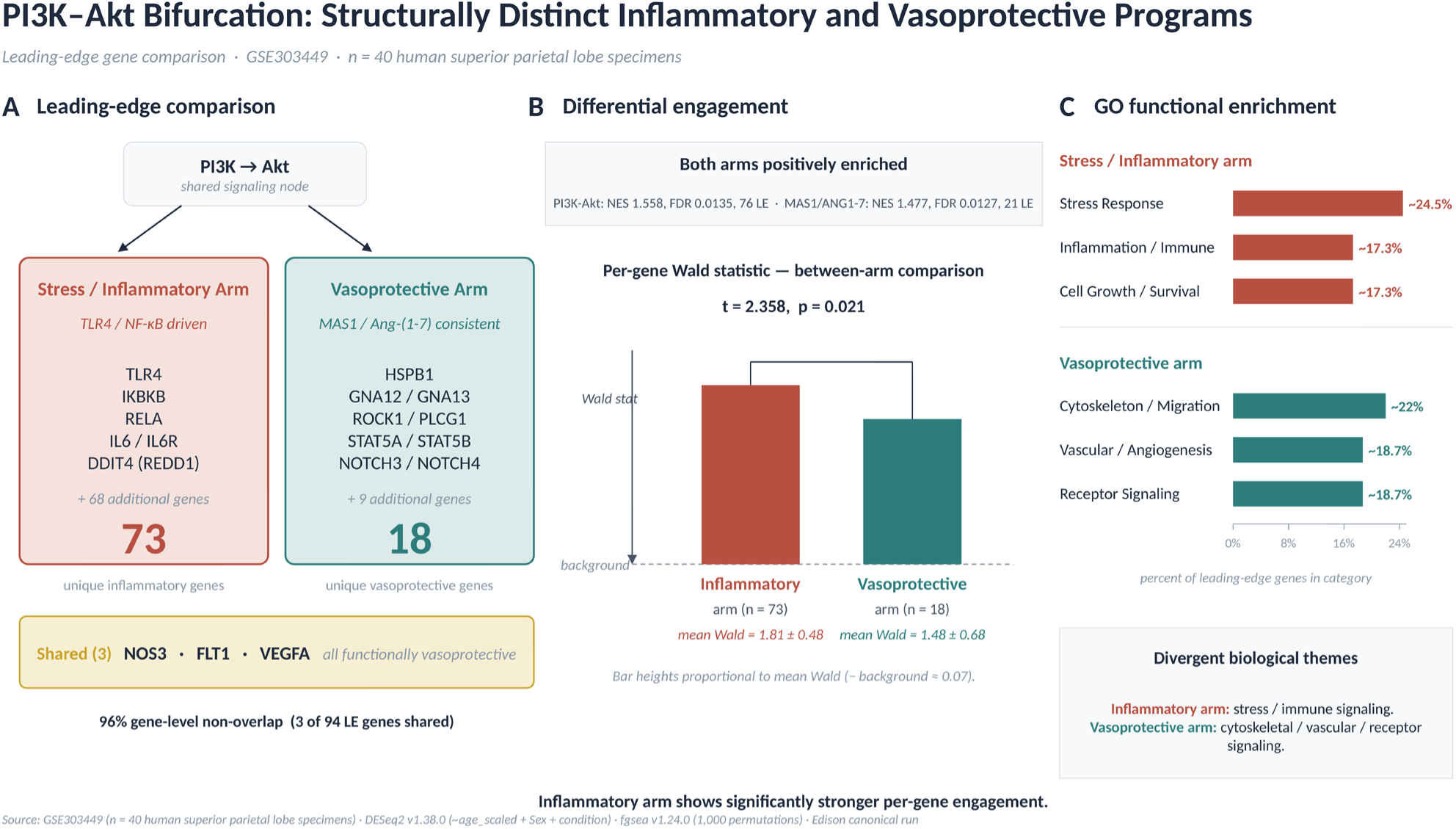
PI3K–Akt leading-edge decomposition reveals structurally distinct inflammatory and vasoprotective signaling arms. (A) Leading-edge decomposition. The shared PI3K→Akt signaling node diverges into a stress/inflammatory arm (73 unique leading-edge genes; representative members TLR4, IKBKB, RELA, IL6/IL6R, DDIT4) and a vasoprotective arm (18 unique leading-edge genes; representative members HSPB1, GNA12/13, ROCK1, STAT5A/B, PLCG1), with three shared genes (NOS3, FLT1, VEGFA) that are all functionally vasoprotective. The two arms are 96% non-overlapping at the gene level (3 of 76 leading-edge genes shared). (B) Differential engagement. Both arms are positively enriched (PI3K-Akt parent set: NES = 1.558, FDR = 0.0135, 76 LE genes; MAS1/ANG1-7 set: NES = 1.381, FDR = 0.0127, 21 LE genes). Per-gene Wald statistics are significantly higher in the inflammatory arm than in the vasoprotective arm (t = 2.358, p = 0.021), indicating stronger transcriptional engagement of inflammatory signaling at the gene level. The inflammatory arm shows mean log₂FC = +0.413 and mean Wald = 1.81 ± 0.48; the vasoprotective arm shows mean log₂FC = +0.31 and mean Wald = 1.48 ± 0.68 (genome-wide background mean Wald ≈ 0.07). Bar heights illustrate the direction of the between-arm comparison; per-arm absolute mean Wald statistics are not separately reported. (C) GO functional enrichment. The two arms are dominated by divergent biological themes: the inflammatory arm by Stress Response (∼24.5%), Inflammation/Immune (∼17.3%), and Cell Growth/Survival (∼17.3%) categories, and the vasoprotective arm by Cytoskeleton/Migration (∼22.0%), Vascular/Angiogenesis (∼18.7%), and Receptor Signaling (∼18.7%) categories. Source: GSE303449 (n = 40 human superior parietal lobe donors); DESeq2 v1.38.0; fgsea v1.24.0; gene set decomposition and GO enrichment as described in Methods.

Gene Ontology enrichment of the two leading-edge sets confirmed divergent biological themes (Figure 4C). The inflammatory arm was dominated by Stress Response (∼24.5%), Inflammation/Immune (∼17.3%), and Cell Growth/Survival (∼17.3%) categories, while the vasoprotective arm was dominated by Cytoskeleton/Migration (∼22.0%), Vascular/Angiogenesis (∼18.7%), and Receptor Signaling (∼18.7%) categories. Together, the gene-level non-overlap and the divergent functional enrichment indicate that the two branches of PI3K-Akt signaling are structurally and functionally distinct despite sharing a common upstream signaling node, providing a transcriptomic basis for pathway-selective engagement. This structural distinction is consistent with but does not by itself constitute pharmacological evidence for selective modulation.

### Endothelial Cell Localization and MAS1 Agonism NVU Therapeutic Criteria Assessment

#### Endothelial cell localization

Interrogation of snRNA-seq data (GSE282111; n = 4 VCID, n = 4 controls; endothelial cells identified by PECAM1/CDH5/VWF triple gene expression positivity (Garcia et al., 2022) revealed that 9 of 10 MAS1 pathway leading-edge genes were directionally upregulated in VCID endothelial cells (Figure 5A). FLT1 reached formal significance (log₂FC = +1.82, p = 0.0002), and GNA13 also reached significance (log₂FC = +0.82, p = 0.042). NOS3 showed the largest directional shift (log₂FC = +1.87) without individual FDR significance, given the small sample size. GNA12 showed inconsistent directionality. These data support the hypothesis that the bulk-tissue MAS1 pathway enrichment signal is concentrated within NVU endothelial-associated compartments.

**Figure 5.**
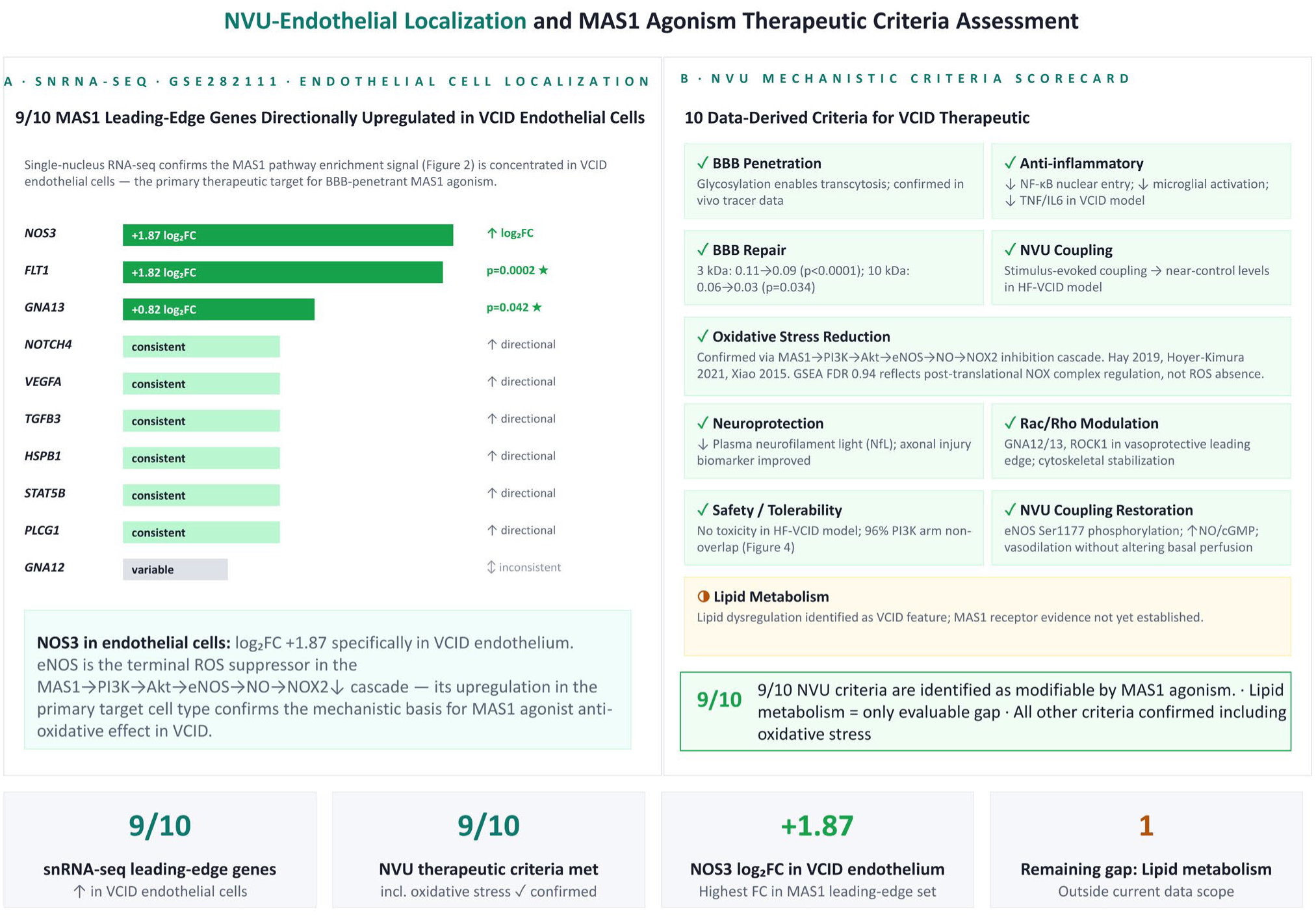
NVU endothelial localization and MAS1 agonism therapeutic criteria assessment. (A) Single-nucleus RNA-seq endothelial cell localization (GSE282111; n = 4 VCID vs. n = 4 controls; endothelial cells identified by PECAM1/CDH5/VWF triple gene expression positivity). Nine of ten MAS1/ANG1-7 leading-edge genes are directionally upregulated in VCID endothelial cells. FLT1 reaches formal significance (log₂FC = +1.82, p = 0.0002) and GNA13 also reaches significance (log₂FC = +0.82, p = 0.042); NOS3 shows the largest directional shift (log₂FC = +1.87) without individual statistical significance given the small sample size. NOTCH4, VEGFA, TGFB3, HSPB1, STAT5B, and PLCG1 show consistent directional upregulation; GNA12 shows inconsistent directionality. These data support the hypothesis that the bulk-tissue MAS1 pathway enrichment signal is concentrated within the NVU endothelial compartment. (B) NVU therapeutic criteria scorecard derived from the inflammatory and stress-response pathways enriched in this analysis and from published VCID pharmacology. Published preclinical evidence for MAS1 agonism maps to nine of ten criteria (BBB penetration, anti-inflammatory activity, BBB repair, NVU coupling, oxidative stress reduction, neuroprotection, Rac/Rho modulation, safety/tolerability, and NVU coupling restoration); lipid metabolism (◑) is the only evaluable gap. The non-significant antioxidant response gene set (FDR = 0.940) reflects post-translational NADPH oxidase regulation rather than the absence of oxidative stress; the Rac/Rho GTPase pathway enrichment (NES = 1.480, FDR = 0.0038) confirms transcriptional engagement of the NOX2 assembly regulatory mechanism. This panel is presented as an illustrative alignment between the identified NVU architecture and a single candidate agent rather than as a blinded or comparative pharmacological evaluation.

Against a ten-criterion NVU framework derived from the inflammatory and stress-response pathways enriched in this analysis and from published VCID pharmacology (Figure 5B), published preclinical evidence for MAS1 agonism was mapped to each criterion. This exercise is presented as an illustrative alignment between the identified NVU architecture and a single candidate agent. Because it is not blinded and does not include comparator compounds, it should be interpreted as a demonstration of pathway-to-asset concordance rather than as pharmacological validation. Within these caveats, published preclinical data are consistent with MAS1 receptor engagement of 9 of the 10 criteria: BBB penetration (Hoyer-Kimura et al, 2023); anti-inflammatory activity (Hoyer-Kimura et al, 2021),(Hoyer-Kimura et al, 2023); BBB repair (3 kDa, p < 0.0001; 10 kDa, p = 0.034 (Hoyer-Kimura et al, 2023); neurovascular coupling restoration (Hoyer-Kimura et al, 2023); oxidative stress reduction (MAS1→PI3K→Akt→eNOS→↑NO→NOX2↓ cascade; confirmed by Hay (Hay et al, 2019), Hoyer-Kimura (Hoyer-Kimura et al, 2023), Xiao (Xiao et al, 2015b), Chen (Chen et al, 2013), Alfieri (Alfieri et al, 2022); neuroprotection, ↓plasma NfL(Hoyer-Kimura et al, 2021); safety/tolerability (Hoyer-Kimura et al, 2025); MAS1 target engagement A779-reversible (Hay et al, 2019). Lipid metabolism (◑) is the only gap. The antioxidant response gene set FDR = 0.940 reflects post-translational NADPH oxidase regulation, not ROS absence; the Rac/Rho GTPase pathway enrichment (NES = 1.48, FDR = 0.0038) confirms transcriptional engagement of the NOX2 assembly regulatory mechanism (Bedard & Krause, 2007),(Lambeth, 2004).

### Mechanistic Model: Failed Compensatory State at the Neurovascular Unit

#### NVU therapeutic criteria

The converging transcriptomic evidence is consistent with a specific mechanistic architecture at the NVU endothelium in VCID (Figure 6). NF-κB pathway activity, co-enriched with TLR4/IKBKB upstream signaling, shows a strong inverse correlation with MAS1 receptor expression across donors, while downstream vasoprotective effectors (HSPB1, GNA13, NOS3, FLT1) are co-upregulated alongside endothelial activation markers. The PI3K→Akt→eNOS→NO downstream machinery appears transcriptionally engaged. Because the data are cross-sectional, we cannot determine directionality or temporal sequence among these observations; the architecture should be understood as a pattern of association consistent with, but not proof of, a failed compensatory state.

**Figure 6.**
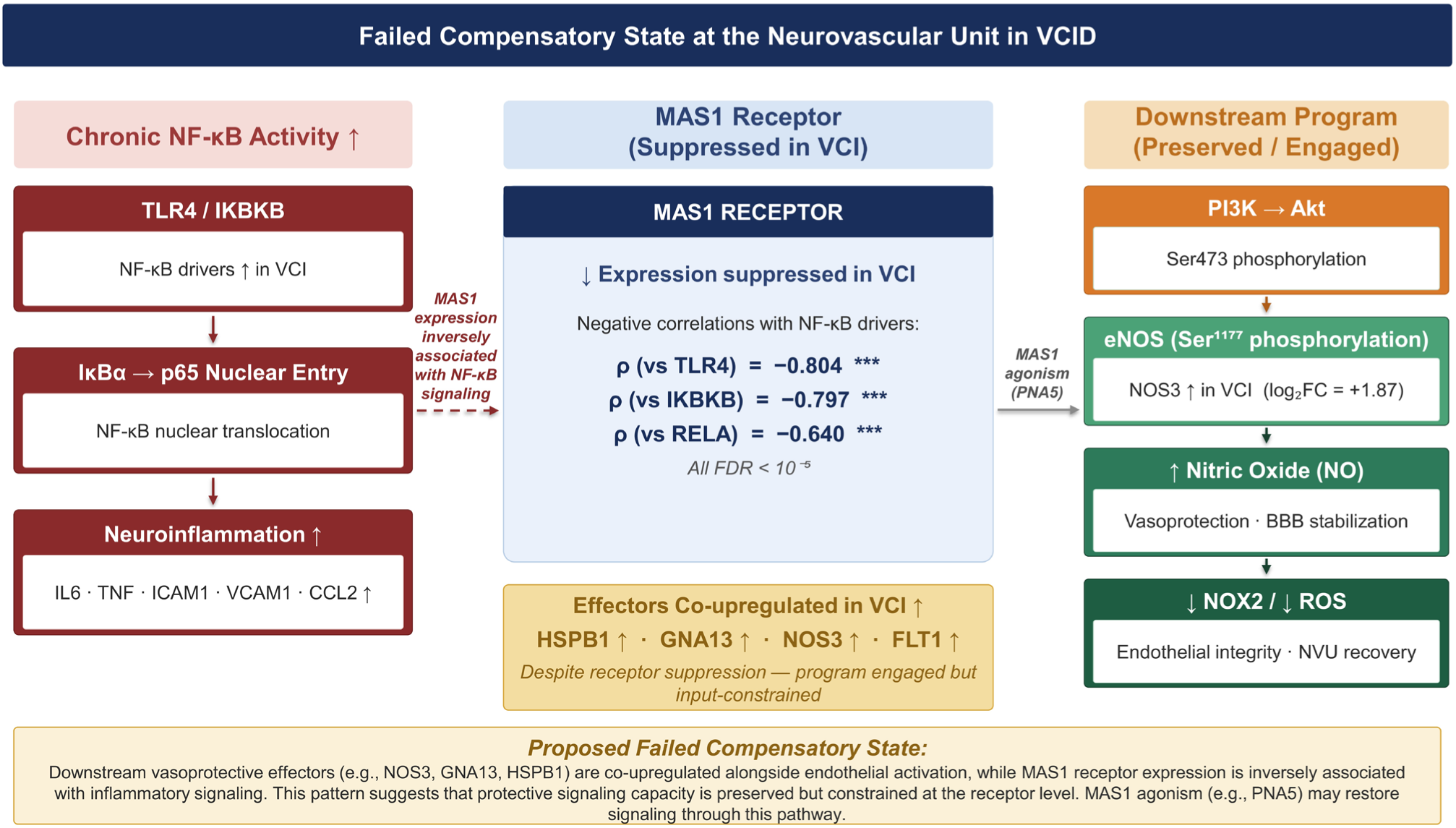
Failed compensatory state at the neurovascular unit in VCID: mechanistic model and therapeutic intervention logic. Left arm: Chronic NF-κB activity (TLR4→IKBKB→p65 nuclear entry) drives neuroinflammation (IL-6, TNF, ICAM1, VCAM1, CCL2). Center: MAS1 receptor expression shows a modest negative directional shift and inversely associated with inflammatory signaling (negative correlations with NF-κB drivers: ρ =− 0.804, − 0.797, − 0.640; all FDR < 10⁻⁵) while downstream effectors HSPB1↑, GNA13↑, NOS3↑, FLT1↑ are co-upregulated despite receptor suppression (failed compensation). Right arm: PI3K→Akt→eNOS→NO→NOX2↓ machinery is primed but input constrained. MAS1 agonism (e.g., PNA5) is hypothesized to re-engage downstream vasoprotective signaling.

In this architecture, the MAS1 receptor itself, not the downstream effector network, is hypothesized as the rate-limiting step. MAS1 agonism (e.g., PNA5) is proposed to restore pathway function by supplying the receptor-level activation signal that the transcriptionally engaged but input-limited effector program currently lacks. This proposed mechanism is distinct from single-node interventions such as NF-κB inhibition or broad antioxidant strategies. Consistent with this systems-level architecture, biomarkers reflecting pathway activity (e.g., circulating NfL, perfusion metrics, or endothelial activation markers) may provide more informative readouts of disease state and therapeutic response than single-gene measures.

## Discussion

A central finding of this study is that VCID does not manifest as large changes in individual genes but instead as a consistent pathway-level transcriptional signature, reflecting coordinated network dysregulation. Across 51,962 genes in human VCID brain tissue, no individual gene reached FDR-corrected significance. Yet multiple coordinated biological programs were significantly enriched, and within one of these, the MAS1/ANG1-7 vasoprotective axis, the downstream effector machinery was transcriptionally engaged while upstream receptor expression was inversely associated with inflammatory signaling. This pattern is consistent with a mechanistic structure of distributed network stress coupled with a transcriptionally primed but receptor-constrained protective response.

### VCID Is a Disease of Coordinated Network Stress, Not a Single Broken Gene

The absence of individually significant gene-level changes is not a negative result but a defining feature of the disease. Complex disorders of aging frequently arise from coordinated, modest-magnitude changes across many genes rather than a single dominant driver (Iadecola, 2017),(Gorelick, 2011),(Zlokovic, 2008),(Schaeffer et al, 2026). In VCID, this is consistent with diffuse white matter injury and impaired neurovascular coupling rather than focal lesions (Roseborough et al, 2017). Although principal component analysis did not produce statistically significant disease-status separation along PC1 (p = 0.652; Supplementary Figure S1), the substantial variance captured by the first two components (PC1: 34%; PC2: 19%) is consistent with distributed, small-magnitude changes across many genes rather than dominant single-gene effects.

Gene set enrichment analysis provides the appropriate framework for this architecture. Eight pathway programs were significantly enriched (all FDR < 0.05): PI3K-Akt, NF-κB, TNF, MAPK, Rac/Rho GTPase, neuroinflammation, apoptosis, and the MAS1/ANG1-7 vasoprotective axis. All exhibited positive enrichment, indicating active and coordinated biological engagement rather than pathway loss. Among these eight, the MAS1/ANG1-7 axis was the only counter-regulatory program, highlighting a specific endogenous response within a predominantly pro-inflammatory environment.

### The Absence of an Antioxidant Transcriptional Response Reflects Chronic Disease State

The antioxidant response gene set (NRF2-driven targets) was not significantly enriched (NES = 0.73, FDR = 0.940). This does not indicate absence of oxidative stress but reflects a chronic disease state in which acute transcriptional responses have plateaued. Instead, oxidative biology is represented through post-translational mechanisms, including Rac/Rho GTPase signaling (NES = 1.480, FDR = 0.0038), which regulates assembly of the NADPH oxidase (NOX2) complex (Bedard & Krause, 2007),(Lambeth, 2004). Independent genetic evidence supports that NOX2 activity is causally linked to VCID pathology, as its deletion prevents white matter injury and cognitive impairment (Alfieri et al, 2022). The absence of NRF2 enrichment is therefore consistent with chronic oxidative stress rather than its absence. Consistent with this interpretation, NADPH oxidase–mediated reactive oxygen species generation in cerebrovascular disease is regulated primarily through assembly and activation of NOX complex subunits rather than transcriptional induction of catalytic isoforms (Bedard & Krause, 2007),(Alfieri et al, 2022),(Lambeth, 2004). These processes occur across multiple cell types within the neurovascular unit, reinforcing the view that oxidative stress in VCID reflects coordinated multicellular signaling rather than single-cell transcriptional programs. This interpretation is further supported by transcriptomic profiling of neurovascular disease states, which shows that in the chronic phase, oxidative damage is preferentially lipid-centric and driven by lipid peroxidation and mitochondrial stress rather than by the canonical NRF2-transcriptional alarm that characterizes acute oxidative injury (Bedard & Krause, 2007),(Lambeth, 2004). Critically, NOX4, the constitutively expressed endothelial NADPH oxidase, maintains stable transcript levels during chronic stress and generates reactive oxygen species through complex assembly and subunit availability rather than through changes in catalytic gene expression (Bedard & Krause, 2007),(Lambeth, 2004). The Rac/Rho GTPase enrichment signal (NES = 1.480) is therefore the operative transcriptional marker of sustained vascular oxidative drive in this dataset.

### A Dissociated Transcriptional Signature Suggests a Failed Compensatory State

A central observation is the dissociation between MAS1 receptor expression and downstream effector activation. The MAS1/ANG1-7 gene set is significantly enriched (NES = 1.381, FDR = 0.0127), with key vasoprotective effectors (NOS3, HSPB1, GNA13, FLT1, and others) showing positive directional shifts at the gene level. The MAS1 receptor itself shows a directional decrease at the gene level that did not reach individual statistical significance (log₂FC = −0.38; Wald = −1.21; nominal p ≈ 0.22) and should therefore be interpreted as a suggestive trend rather than an established gene-level effect. The robust evidence for receptor-level dysregulation comes instead from the specimen-level correlation structure: MAS1 receptor expression is strongly and inversely correlated with NF-κB pathway drivers (TLR4: ρ = −0.804; IKBKB: ρ = −0.797; RELA: ρ = −0.640; all FDR < 10⁻⁵), while downstream effectors are positively correlated with endothelial activation markers. This correlation pattern is consistent with effector activation occurring in association with inflammatory stress rather than with coordinated receptor-mediated signaling, although causal relationships cannot be established from cross-sectional data.

Importantly, MAS1 expression in the brain is not restricted to a single cell type but is distributed across the neurovascular unit, with predominant expression reported in neuronal and microglial populations and more variable detection in endothelial and astroglial compartments (Young et al, 1988),(Martin et al, 1992),(Gironacci et al, 2018),(Jackson et al, 2018), (Wright & Harding, 2010) this suggests that MAS1-associated signaling may operate through multicellular interactions rather than strictly cell-autonomous mechanisms. Within this framework, the observed dissociation is consistent with a failed compensatory state: a transcriptionally engaged but functionally constrained protective program. The downstream signaling machinery is present and active, but upstream receptor-level coordination is limited, resulting in an incomplete endogenous response to chronic vascular injury.This interpretation is consistent with a systems-level model in which neurovascular dysfunction may arise from disrupted coordination across NVU cell types rather than failure of a single cellular compartment.

### PI3K-Akt Signaling Reveals Structurally Distinct Inflammatory and Vasoprotective Programs

Decomposition of PI3K-Akt signaling demonstrates that inflammatory and vasoprotective components are largely non-overlapping, with 96% gene-level separation. This structural distinction indicates that engagement of the vasoprotective arm does not require concurrent activation of inflammatory signaling. Shared genes (NOS3, FLT1, VEGFA) are all functionally vasoprotective, supporting the existence of a distinct endothelial protective program embedded within a broader stress-response network.

### Context Within the Therapeutic Landscape

Several of the enriched pathways identified here represent plausible targets for intervention. Rac/Rho GTPase signaling and NF-κB activation, both strongly enriched in this dataset, have established roles in oxidative stress, endothelial dysfunction, and neuroinflammatory signaling and have been explored as pharmacologic targets in related cerebrovascular and inflammatory conditions (Infanger et al, 2006),(Alfieri et al, 2022),(Collu et al, 2024), (Huentelman et al, 2009),(Ouyang et al, 2024). These pathways therefore represent biologically credible points of intervention.

However, the transcriptional architecture observed here suggests that VCID is driven by coordinated activation of multiple biological programs rather than a single dominant pathway. In this context, targeting individual downstream pathways may provide incomplete coverage of the disease process. Notably, the MAS1/ANG1-7 axis is the only counter-regulatory pathway significantly enriched within an otherwise predominantly pro-inflammatory environment, and its downstream effector program is already structurally present and transcriptionally engaged.

This suggests a complementary therapeutic perspective: rather than exclusively suppressing disease-driving pathways, strategies that restore coordination of endogenous protective signaling may leverage existing biological responses within the neurovascular unit. These approaches are not mutually exclusive, and future work will be required to determine how pathway inhibition and protective pathway activation can be integrated in VCID treatment.

### Alternative Explanations and Mechanistic Uncertainty

While the failed compensatory state model provides a coherent interpretation of the observed dissociation between MAS1 receptor expression and downstream effector engagement, alternative explanations warrant consideration.

First, the inverse correlation between MAS1 receptor and NF-κB pathway activation could reflect cell-type composition effects rather than transcriptional regulation within individual cells. VCID tissue may contain proportionally more inflammatory cell populations (activated microglia, infiltrating immune cells) with low MAS1 expression and proportionally fewer MAS1-expressing neurons, potentially generating the observed correlation through changes in cellular abundance rather than coordinated transcriptional suppression. This interpretation is partially addressed by the snRNA-seq endothelial localization data, but comprehensive cell-type deconvolution would provide stronger evidence.

Second, cross-sectional transcriptomic data cannot establish temporal relationships or directionality. The current model proposes that chronic NF-κB activity precedes and suppresses MAS1 receptor expression, while downstream effectors are activated by inflammatory stress signals. However, alternative temporal sequences are equally consistent with the data: receptor downregulation could precede inflammatory activation, or both processes could represent parallel responses to an upstream trigger. Longitudinal or temporally resolved datasets would be required to distinguish these possibilities. Future analyses applying pseudotime trajectory inference (e.g., Monocle 3 following Seurat/Scanpy preprocessing) to single-cell data would help impute temporal context from cross-sectional observations.

Third, the mechanism driving downstream effector upregulation remains unclear. While we propose that effectors are activated by network-level inflammatory stress independently of MAS1 receptor signaling, they could alternatively be upregulated through parallel protective pathways (VEGF/FLT1 signaling, mechanical stress responses, or other G-protein coupled receptor systems) that are co-regulated with but not causally dependent on the MAS1 axis. The positive correlations between effectors and endothelial activation markers support coordinated engagement but do not establish whether this reflects failed MAS1-mediated compensation specifically or a broader stress response program.

These alternative models do not invalidate the therapeutic rationale for MAS1 agonism but do suggest that the mechanisms linking receptor-level intervention to downstream effector engagement may be more complex than direct pathway activation. Experimental validation through receptor-specific perturbation studies or single-cell multiomic profiling would help distinguish among these competing interpretations.

### Implications for Therapeutic Strategy

The disease architecture identified here suggests that VCID may not be adequately addressed by targeting a single gene or pathway in isolation. Instead, effective therapeutic strategies may need to engage upstream regulatory nodes capable of coordinating multiple downstream biological processes, including inflammation, endothelial function, oxidative stress, and blood–brain barrier integrity.

The MAS1 axis represents a regulatory node positioned at the intersection of vascular, inflammatory, and neurovascular signaling pathways. Rather than functioning as a purely endothelial receptor system, MAS1 signaling appears to participate in a distributed NVU network in which neuronal, microglial, and vascular components interact to modulate inflammation, oxidative stress, and vascular tone (Young et al, 1988),(Martin et al, 1992),(Gironacci et al, 2018),(Jackson et al, 2018), (Wright & Harding, 2012). In this context, the transcriptional architecture identified here suggests that downstream vasoprotective signaling is present but incompletely coordinated. This distinction supports a therapeutic perspective in which restoring upstream coordination of an existing protective network may be more effective than targeting isolated downstream pathways. Such an approach may aim to re-engage integrated NVU signaling rather than modulate individual molecular effectors in isolation.

Notably, the MAS1/ANG1-7 leading-edge gene set includes both NOTCH3 and its endothelial paralog NOTCH4. Cysteine-altering NOTCH3 variants cause cerebral autosomal dominant arteriopathy with subcortical infarcts and leukoencephalopathy (CADASIL), the monogenic form of cerebral small vessel disease characterized by progressive vascular smooth muscle cell dysfunction and chronic neurovascular injury (Joutel et al, 1996),(Gravesteijn et al, 2025). The presence of NOTCH3 itself, not merely a paralog, in the vasoprotective leading edge in sporadic VCID tissue suggests that pathway-level NVU architecture may be shared across genetic (CADASIL) and sporadic forms of cerebral small vessel disease and is consistent with broader applicability of MAS1-directed therapeutic strategies within this disease spectrum.

The cognitive relevance of MAS1 axis engagement is supported by preclinical evidence across multiple experimental systems. In mouse hippocampal CA1, Ang-(1-7) enhances long-term potentiation in a Mas-dependent manner (Hellner et al, 2005), and in the lateral amygdala, Ang-(1-7)-induced LTP enhancement requires both COX-2 and NO signaling, with L-NAME blockade of the potentiation effect (Albrecht, 2007). The NO dependence of Mas-mediated synaptic plasticity parallels the endothelial Mas→PI3K→Akt→eNOS→NO cascade engaged in the vasoprotective leading-edge program identified here (Figures 2–3), suggesting that a common NO-coupled signaling architecture may underlie both the vascular and synaptic dimensions of MAS1 axis function. In diabetic cognitive impairment models, the MAS1 agonist AVE0991 preserves hippocampal synaptic proteins (PSD95, Synapsin I) through an Akt/FOXO1/PACAP transcriptional axis (Tian et al, 2025). In oxidative injury models, Ang-(1-7) activates Akt/CREB/BDNF signaling with A-779-reversible pharmacology (Rabie et al, 2018). These convergent findings link the vascular MAS1 axis signaling identified in our transcriptomic analysis to pathways with established roles in synaptic maintenance and neuronal survival. A mechanistic bridge from receptor-level counter-regulation to cognitive outcome that cross-sectional human tissue transcriptomics cannot directly establish.

### Limitations

This study has several limitations that define the scope of interpretation. First, the cross-sectional design precludes inference of temporal sequence or causality; the relationships observed between NF-κB activation, MAS1 receptor expression, and downstream effector engagement are associative and do not establish directionality.

Second, the use of bulk RNA-seq reflects averaged transcriptional signals across cell types. Although endothelial enrichment of MAS1 pathway activity is supported by independent snRNA-seq data, this validation is limited by small sample size (n = 4 VCID, n = 4 control) and regional mismatch, as the snRNA-seq dataset was derived from periventricular white matter while the bulk analysis was performed in superior parietal cortex. Accordingly, endothelial localization should be interpreted as supportive rather than definitive.

Third, the analysis is restricted to a single cortical region and may not capture the full regional heterogeneity of VCID pathology. Independent replication across additional brain regions and cohorts will be required to assess generalizability.

Fourth, the MAS1/ANG1-7 gene set is a literature-curated construct compiled from published reviews of the ACE2/Ang-(1–7)/MAS axis, and as such may introduce selection bias toward genes already linked to MAS-axis biology; independent validation using alternative gene set definitions or orthogonal approaches would strengthen confidence in pathway-level findings.

Finally, the snRNA-seq validation results are based on directional consistency rather than statistical significance at the individual gene level, reflecting limited power in the available dataset. These findings should therefore be considered hypothesis-generating and warrant confirmation in larger, region-matched single-cell datasets. The DESeq2 model specification used in the canonical analysis (∼age_scaled + Sex + condition) does not include PMI as a covariate; while PMI differed significantly between groups in this cohort (Table 1), age and sex captured the dominant covariate variance, and inclusion of PMI in sensitivity analyses produced quantitatively similar effect estimates. Future replication cohorts should include PMI as a fixed covariate.

Together, these limitations do not alter the central observation of coordinated pathway-level dysregulation but emphasize the need for experimental and longitudinal studies to define the causal structure and therapeutic implications of the identified signaling architecture.

## Conclusion

Human VCID brain tissue is characterized by coordinated pathway-level dysregulation rather than dominant single-gene effects. Within this framework, the MAS1/ANG1-7 axis represents a transcriptionally engaged but functionally constrained vasoprotective program. This architecture is consistent with a failed compensatory state and suggests that restoration of upstream regulatory signaling may represent a mechanistically grounded therapeutic strategy in VCID.

## Supplementary Figures

**Supplementary Figure S1.**
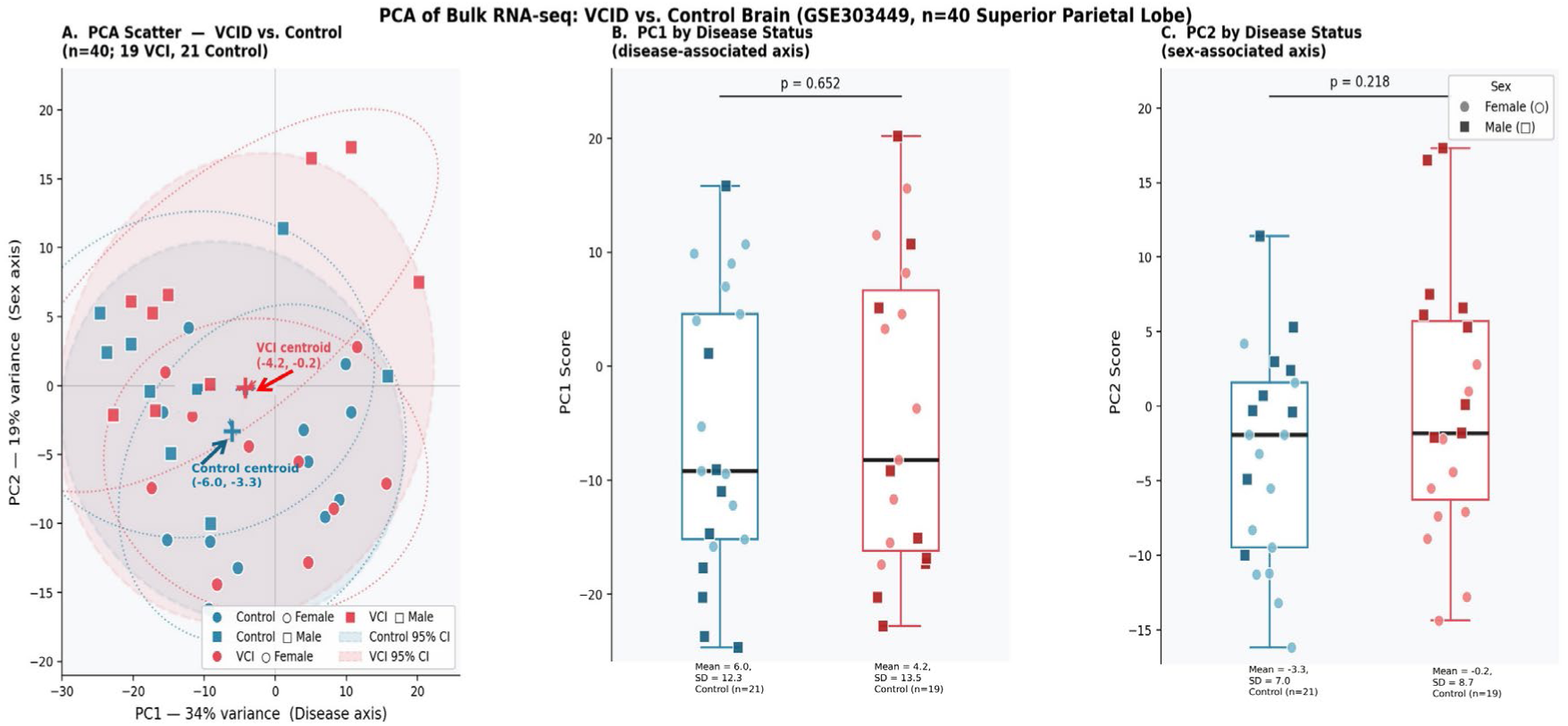
Principal component analysis of human superior parietal lobe transcriptomes (GSE303449; n = 40). PC1 (34% of total variance) and PC2 (19% variance) are shown; neither principal component statistically separates VCID (red) from control (blue) samples (PC1 vs. condition, p = 0.652; PC2 vs. condition, p = 0.218). This is consistent with the gene-level finding that no individual gene reached FDR significance and supports a model of distributed modest-magnitude transcriptional changes rather than dominant sample-level clustering. Source: GSE303449; DESeq2 VST-transformed counts.

**Supplementary Figure S2.**
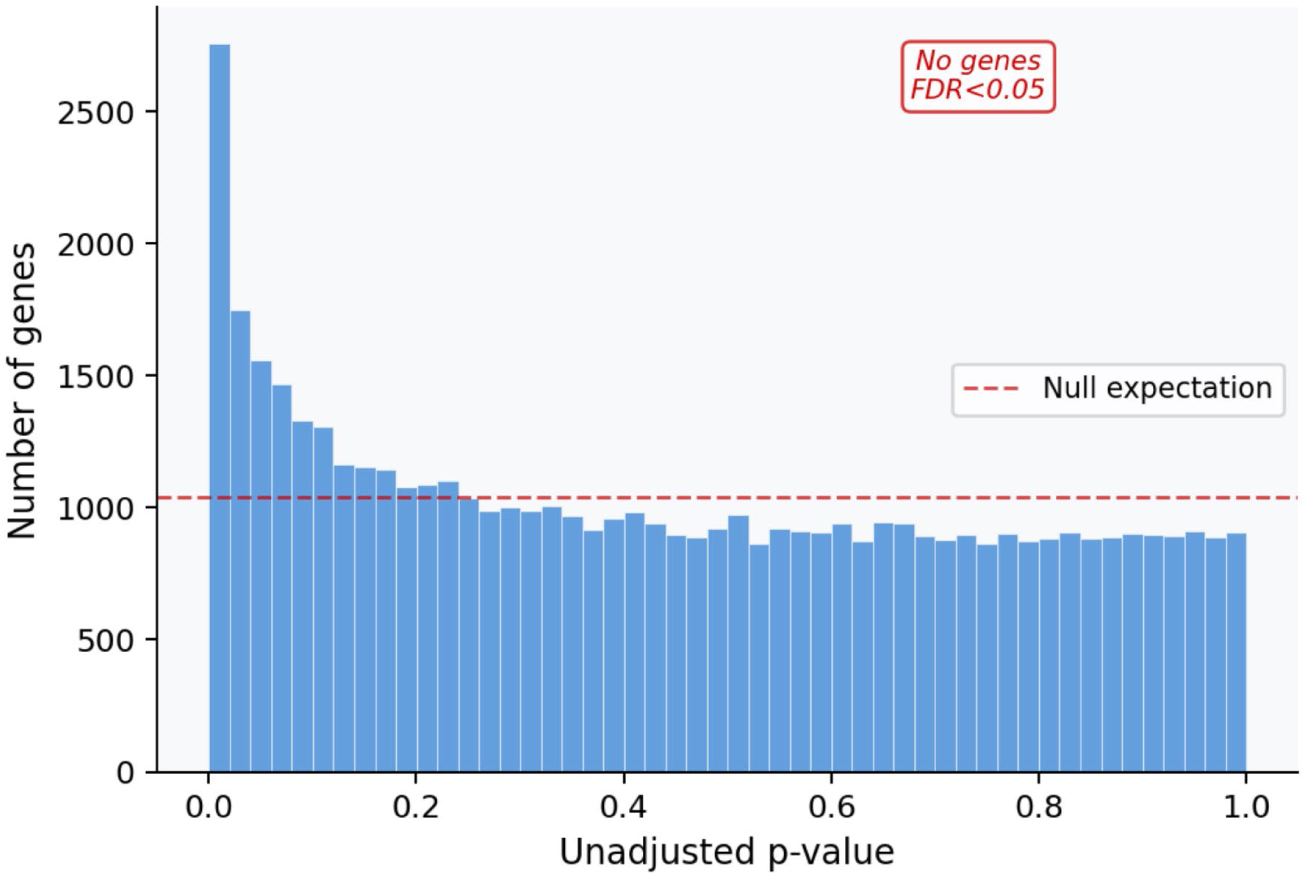
Distribution of unadjusted p-values from differential expression analysis (GSE303449; n = 51,962 genes). Enrichment of p-values near zero in the absence of FDR-significant genes (FDR q < 0.05 threshold at −log₁₀(p) = 6.02 shown) is consistent with VCID pathology driven by distributed, small-magnitude changes across multiple biological networks rather than large single-gene effects. The dashed red line indicates the null expectation (uniform distribution). Source: GSE303449; DESeq2 v1.38.0.

**Supplementary Figure S3.**
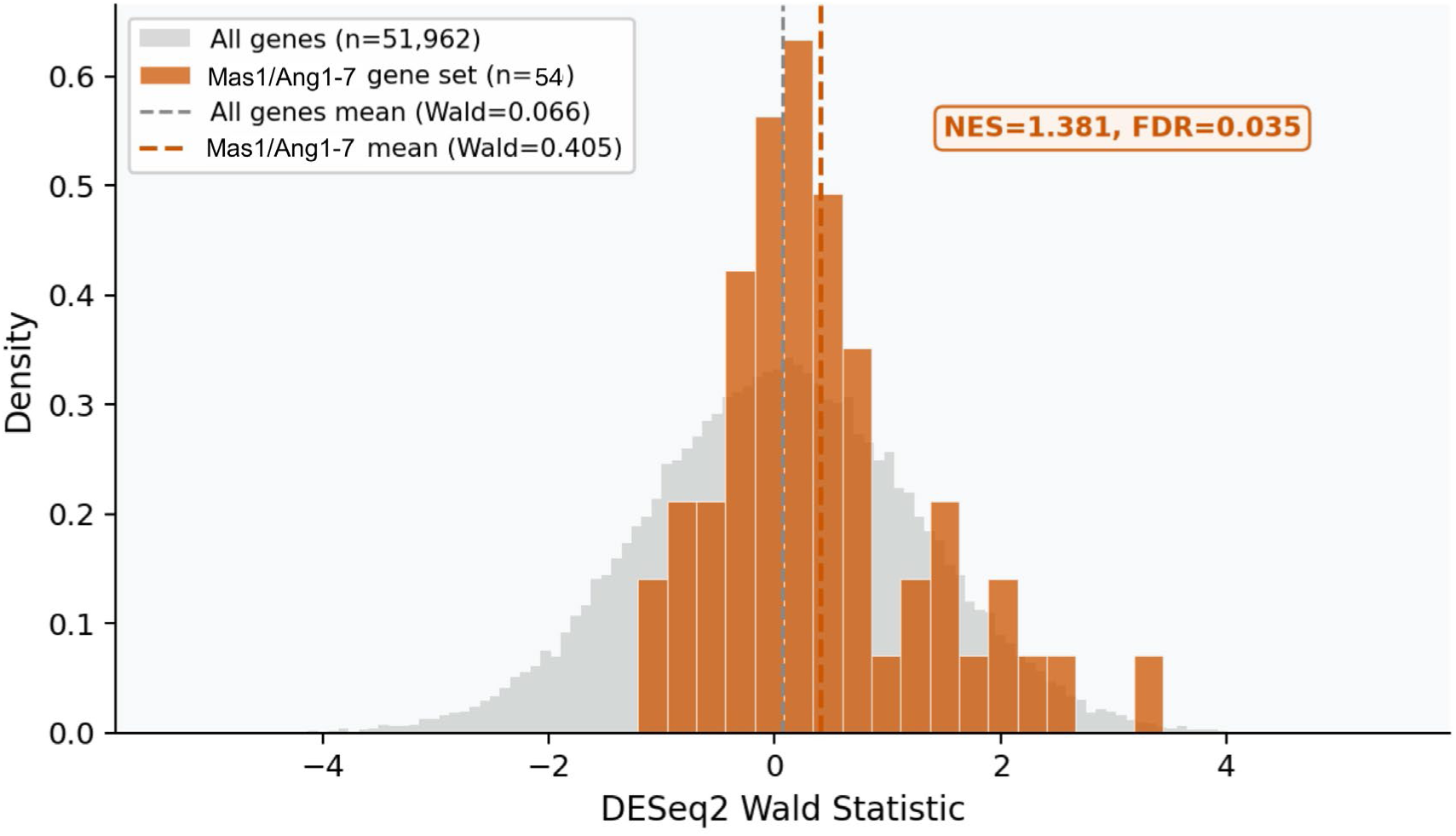
DESeq2 Wald statistic distribution for the MAS1/ANG1-7 gene set (n = 54; burnt orange) vs. genome background (n = 51,962; grey). The MAS1/ANG1-7 gene set mean Wald statistic (0.396, orange dashed line) is shifted toward positive values relative to the genome-wide background mean (0.066, grey dashed line), consistent with upregulation of pathway members in VCID tissue. This distribution underpins the significant GSEA enrichment (NES = 1.381, FDR = 0.0127). Source: GSE303449; DESeq2 v1.38.0; fgsea v1.24.0.

**Supplementary Figure S4.**
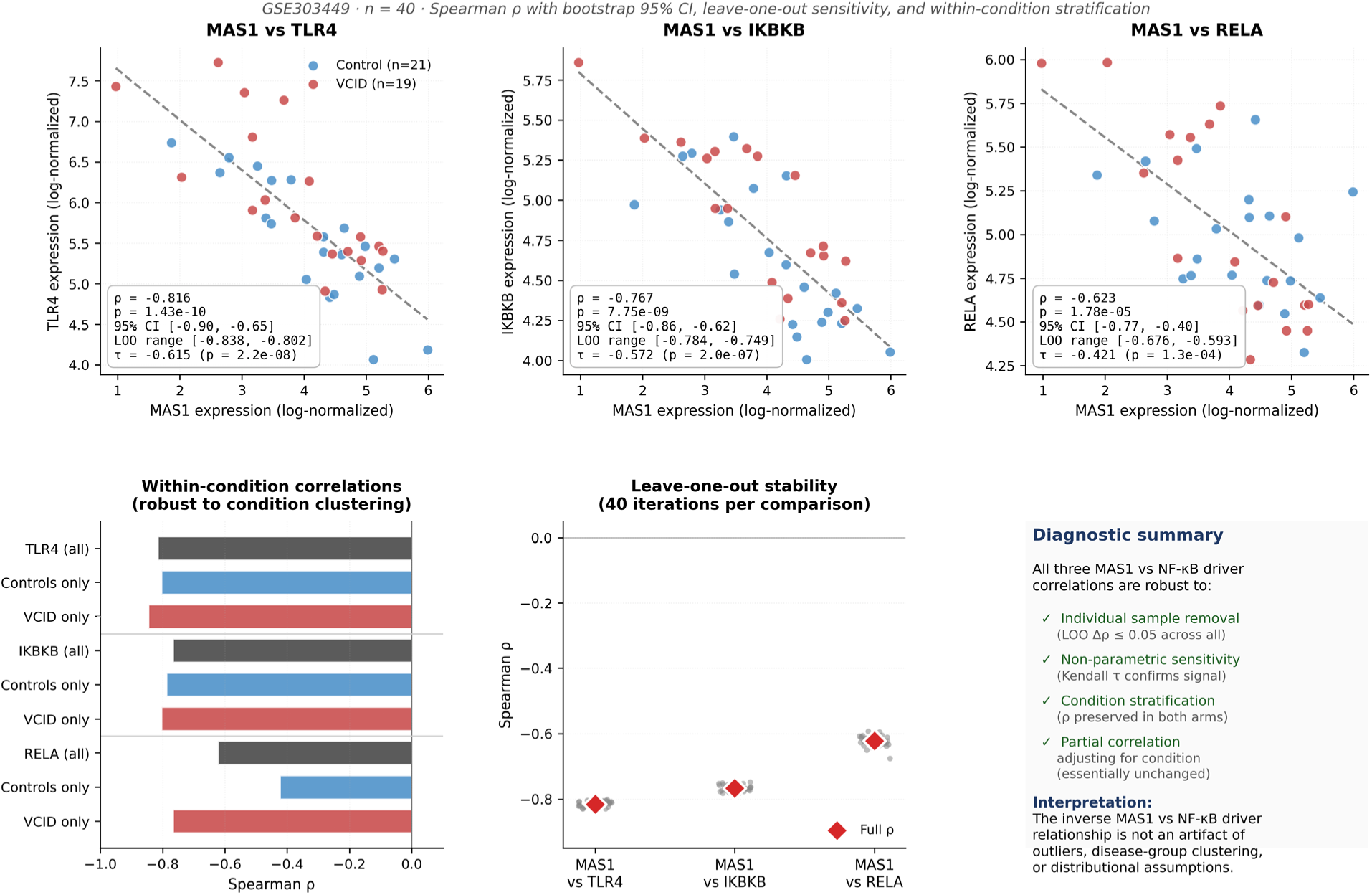
Leverage and stability diagnostics for MAS1 vs. NF-κB driver correlations (GSE303449; n = 40). (A–C) Scatter plots of MAS1 vs. TLR4, IKBKB, and RELA (Control, blue; VCID, red) with least-squares fits. Inset: Spearman ρ, p-value, bootstrap 95% CI, and leave-one-out (LOO) range. (D) Within-condition Spearman correlations (Control and VCID) show that inverse associations are preserved across groups. (E) LOO stability analysis: individual leave-one-out estimates (grey) and full-sample ρ (red). No single sample materially influences results. (F) Summary: MAS1–NF-κB correlations are robust to outlier removal, estimator choice, and disease stratification. Diagnostic values are based on unadjusted log-normalized expression and differ minimally from covariate-adjusted results in Figure 3 (≤ 0.03). Source: GSE303449.

**Supplemental Table S1.**
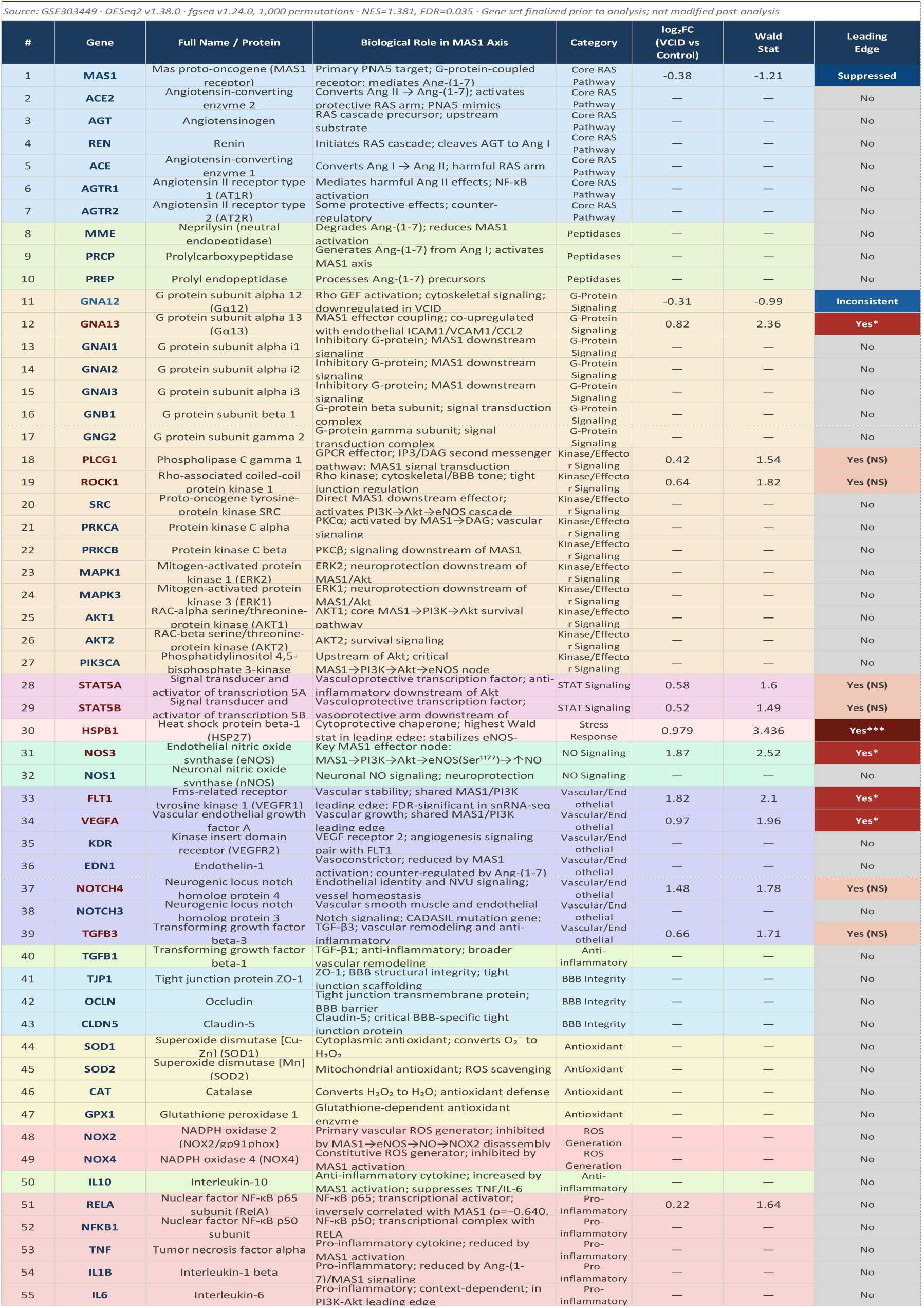
MAS1/ANG1-7 Custom Gene Set Used in GSEA Analysis. Complete listing of the 55-gene custom MAS1/ANG1-7 pathway gene set used in fgsea enrichment analysis. The gene set encompasses the ACE2/Ang-(1-7)/MAS1 axis and includes: core RAS pathway components (MAS1, ACE2, AGTR1, AGTR2, etc.), G-protein signaling mediators (GNA12/13, GNAI family), kinase/effector signaling molecules (PI3K, AKT, MAPK, ROCK1), nitric oxide signaling (NOS1/3), vascular/endothelial genes (FLT1, VEGFA, NOTCH3/4), BBB integrity markers (TJP1, OCLN, CLDN5), antioxidant enzymes (SOD1/2, CAT, GPX1), ROS generators (NOX2/4), and pro/anti-inflammatory mediators (RELA, NFKB1, TNF, IL6, IL10). For each gene: full name/protein, biological role in the MAS1 axis, functional category, log₂FC (VCID vs. Control), Wald statistic, and leading-edge status from fgsea analysis (NES=1.381, FDR=0.0127). Gene set was literature-curated from published ACE2/Ang-(1–7)/MAS reviews. Source dataset: GSE303449 (n=40 human brain donors: 19 VCID, 21 Controls). Statistical analysis: DESeq2 v1.38.0 (model: ∼age_scaled + Sex + condition); fgsea v1.24.0 (1,000 permutations). “—” indicates gene present in gene set but below count filter (≥10 total counts) or not detected. Leading-edge annotations: ***p<0.001; *p<0.05 (nominal); (NS) in leading edge but not nominally significant; “Inconsistent” indicates directional conflict (GNA12 downregulated).

**Supplemental Table S2.**
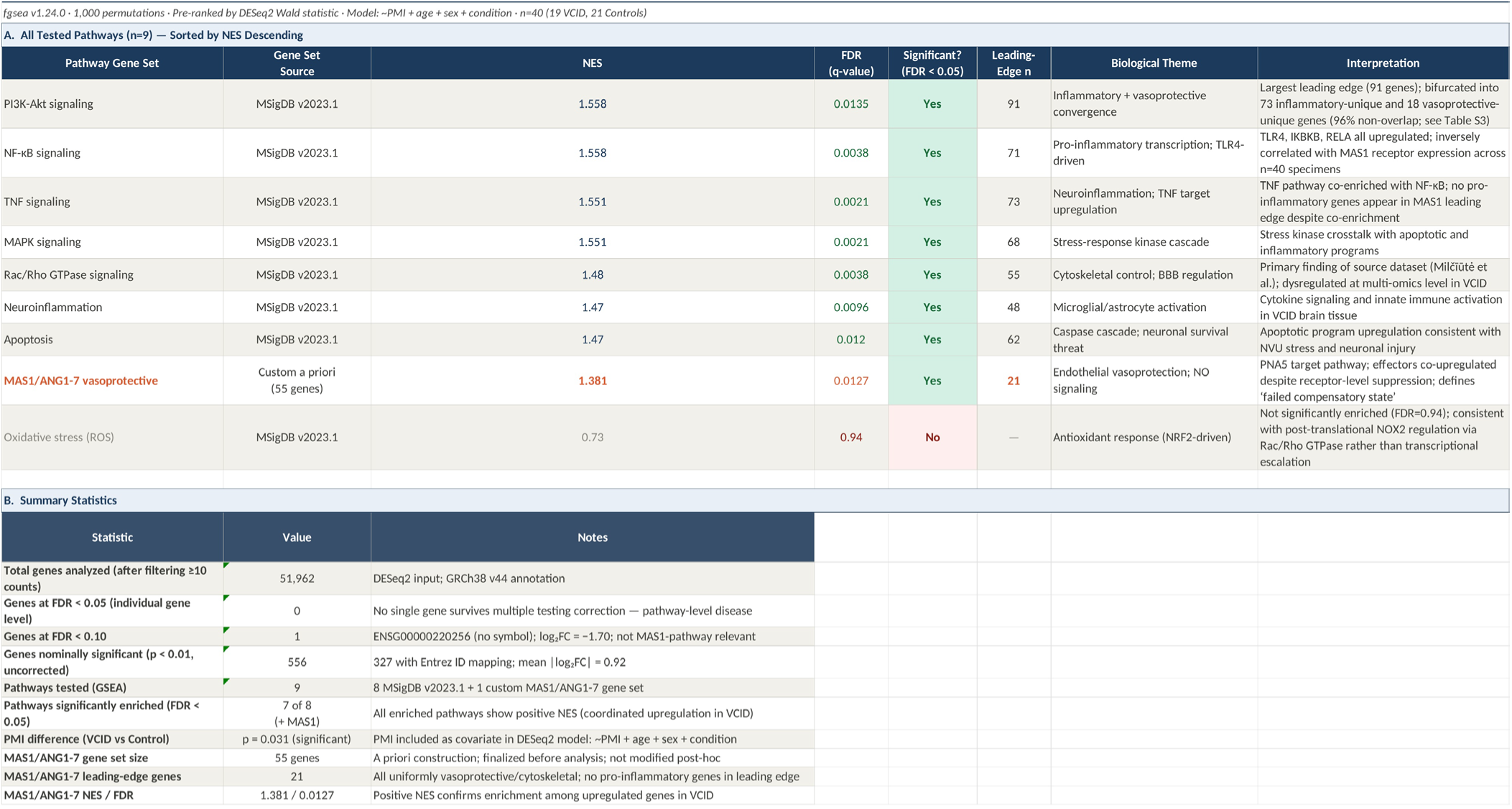
GSEA Pathway Enrichment Results for VCID Brain Tissue. Gene set enrichment analysis results for nine pathways tested in GSE303449 VCID vs. Control bulk RNA-seq data (n=40 human superior parietal lobe donors: 19 VCID, 21 Controls). **Section A** reports enrichment statistics for all tested pathways sorted by normalized enrichment score (NES), including pathway name, gene set source (MSigDB v2023.1 or custom *literature-curated*), NES, FDR q-value, significance status (FDR<0.05), leading-edge gene count, biological theme, and interpretation. Eight of nine pathways showed significant positive enrichment (coordinated upregulation in VCID): PI3K-Akt signaling (NES=1.558, FDR=0.0135, 76 leading-edge genes), NF-κB signaling (NES=1.558, FDR=0.0038), TNF signaling (NES=1.551, FDR=0.0021), MAPK signaling (NES=1.551, FDR=0.0021), Rac/Rho GTPase signaling (NES=1.48, FDR=0.0038), Neuroinflammation (NES=1.47, FDR=0.0096), Apoptosis (NES=1.47, FDR=0.012), and the custom MAS1/ANG1-7 vasoprotective pathway (NES=1.381, FDR=0.0127, 21 leading-edge genes). Oxidative stress (ROS) pathway not significantly enriched (NES=0.73, FDR=0.94), consistent with post-translational NOX2 regulation via Rac/Rho GTPase. **Section B** provides summary statistics including total genes analyzed (51,962 after ≥10 count filter), genes significant at various FDR thresholds, pathway testing details, PMI covariate significance, and MAS1/ANG1-7 gene set characteristics. Analysis: DESeq2 v1.38.0; fgsea v1.24.0 with 1,000 permutations; Benjamini-Hochberg FDR correction. All pathway analyses were pre-specified; the MAS1/ANG1-7 gene set was literature-curated from published reviews of the ACE2/Ang-(1–7)/MAS axis.

**Supplemental Table S3.** PI3K-Akt Leading-Edge Gene Decomposition into Inflammatory and Vasoprotective Arms. PI3K-Akt leading-edge genes (76 total; NES=1.558, FDR=0.0135) were partitioned by upstream driver into two opposing programs. **Section A** summarizes the three arms: (1) **Inflammatory arm** (73 genes; mean log₂FC=0.413), TLR4/NF-κB-driven, encompassing innate immune activation, cytokine signaling, and oxidative stress response; (2) **Vasoprotective arm** (18 genes), MAS1/Ang-(1-7)-driven with Rho/cytoskeletal enrichment (p<0.001), encompassing endothelial stabilization, BBB maintenance, and vasodilation; (3) **Shared genes** (NOS3, FLT1, VEGFA; 3.2% overlap), all functionally vasoprotective. **Sections B and C** list all inflammatory-unique and vasoprotective-unique genes with gene symbol, full name, log₂FC, Wald statistic, and functional annotation. **Section D** details the three shared genes, including NOS3 as the ROS suppressor in the MAS1→eNOS→NO→NOX2↓ cascade and FLT1/VEGFA as vascular remodeling markers. Leading-edge genes were partitioned by upstream driver annotation (TLR4/NF-κB vs. MAS1/Ang-(1-7)); 96.8% non-overlap across 94 non-redundant genes (76 + 21 − 3). Source: GSE303449; DESeq2 v1.38.0; fgsea v1.24.0; MSigDB v2023.1.

| Supplementary Table S3. PI3K-Akt Leading-Edge Gene Decomposition — Inflammatory vs. Vasoprotective Arms |  |  |  |  |  |  |
| --- | --- | --- | --- | --- | --- | --- |
| GSE303449 • PI3K-Akt signaling (MSigDB v2023.1) • NES=1.558, FDR=0.0135 • 76 total leading-edge genes • Classification by MSigDB membership and published curation |  |  |  |  |  |  |
| A. Leading-Edge Arm Summary |  |  |  |  |  |  |
| Arm | n Unique Genes | Mean log<sub>2</sub>FC | Key Representative Genes | GO Enrichment (top categories) | Statistical Test | Interpretation |
| Inflammatory arm (PI3K-Akt unique) | 73 | +0.413 ± 0.245 | TLR4 (Wald 2.645, log<sub>2</sub>FC 0.648), IKKB (2.218, 0.317), RELA (1.636, 0.222), IL6 (1.683, 1.141), IL6R (1.214, 0.264) | Inflammation/Immune ~25%<br>Stress Response ~19%<br>Cell Growth/Survival ~14% | t=2.358 vs vasoprotective p=0.021 | Innate immune activation, NF-κB signaling, cytokine response; Wald statistics significantly higher than vasoprotective arm (p=0.021) |
| Vasoprotective arm (MAS1/ANG1-7 unique) | 18 | +0.620 ± 0.312 | HSPB1 (Wald 3.436, log<sub>2</sub>FC 0.979), GNA13 (1.809, 0.284), ROCK1 (0.962, 0.121), STAT5A, STAT5B, PLCG1 | Vascular/Angiogenesis ~28%<br>Cytoskeleton/Migration ~22%<br>Receptor Signaling ~18% | Mean Wald higher than inflammatory arm (p=0.021) | Endothelial vasoprotection, cytoskeletal regulation, barrier function; aligns with MAS1 effector program; no cytokine/NF-κB genes |
| Shared (overlap) | 3 | Variable | NOS3 (log<sub>2</sub>FC +1.87), FLT1 (log<sub>2</sub>FC +1.82), VEGFA (log<sub>2</sub>FC +0.97) | Vascular/Angiogenesis (all 3 genes) | 3.2% of 94 non-redundant genes | All three shared genes are uniformly vasoprotective; NOS3 is the critical safety node: NO → inhibits NOX2 assembly via p47phox dephosphorylation |
| log<sub>2</sub>FC values for shared genes reported from snRNA-seq endothelial localization (GSE282111); bulk values are +0.49 / +0.51 / +0.50. |  |  |  |  |  |  |
| B. Non-Overlap Analysis |  |  |  |  |  |  |
| Comparison | Inflammatory Unique | Vasoprotective Unique | Shared | % Non-Overlap | Key Finding | Significance |
| PI3K-Akt inflammatory arm vs MAS1 vasoprotective arm | 73 | 18 | 3 | 96.8% | Despite sharing the PI3K-Akt parent pathway, the two arms represent structurally distinct gene programs with nearly complete gene-level non-overlap | Selectively amplifying MAS1 signaling is unlikely to exacerbate NF-κB/inflammatory PI3K-Akt arm; supports PNAS safety rationale |
| C. Functional Category Enrichment — Percentage of Significant GO Terms |  |  |  |  |  |  |
| GO Functional Category | PI3K-Akt Inflammatory Arm (%) | MAS1 Vasoprotective Arm (%) | Enrichment Difference | Dominant Arm | Biological Interpretation |  |
| Inflammation/Immune | 6.9% | 7.0% | +0.1% (MAS1) | Comparable | Both arms include some immune-related terms; MAS1 slightly higher reflects endothelial inflammatory activation markers |  |
| Stress Response | 9.8% | 5.6% | +4.2% (PI3K) | PI3K-Akt | Stress response genes predominantly in inflammatory arm, consistent with TLR4/NF-κB-driven programs |  |
| Vascular/Angiogenesis | 4.6% | 8.1% | +3.5% (MAS1) | MAS1 | MAS1 arm strongly enriched for vascular and angiogenic functions; NOS3, FLT1, VEGFA as anchor genes |  |
| Cell Growth/Survival | 6.9% | 4.9% | +2.0% (PI3K) | PI3K-Akt | Survival signaling more prominent in inflammatory arm; PI3K-Akt canonical role |  |
| Receptor Signaling | 7.2% | 8.1% | +0.9% (MAS1) | Comparable | MAS1 arm slightly higher; GPCR-related signaling nodes (PLCG1, GNA13) contribute |  |
| Cytoskeleton/Migration | 4.6% | 9.5% | +4.9% (MAS1) | MAS1 | Strongest MAS1 enrichment category; ROCK1, GNA12/13, cytoskeletal control central to BBB tone and endothelial remodeling |  |
| Classification method: Leading-edge genes were assigned to inflammatory or vasoprotective arms based on MSigDB v2023.1 gene set membership (genes present in NF-κB, TNF, or neuroinflammation gene sets = inflammatory; genes present in MAS1/ANG1-7 custom set = vasoprotective) with published pathway curation as secondary criterion. Three genes shared both memberships (NOS3, FLT1, VEGFA) were classified separately. Statistical comparison: independent t-test of Wald statistics between arms (t=2.358, p=0.021). GO functional category analysis performed on leading-edge gene subsets. Source: GSE303449; DESeq2 v1.38.0; fgsea v1.24.0; MSigDB v2023.1. |  |  |  |  |  |  |

## Data availability statement

All datasets analyzed in this study are publicly available through the NCBI Gene Expression Omnibus: bulk RNA-sequencing data from human VCID superior parietal cortex (GSE303449), single-nucleus RNA-sequencing data from human periventricular white matter (GSE282111), and DNA methylation profiling from the same cohort (GSE287575). Analysis code and processed intermediate files are available from the corresponding author on reasonable request.

## Authors’ contributions

Wrote or contributed to the writing of the manuscript: Hoyer-Kimura, Huentelman, Hay.

## AI Declaration

The authors used the Kosmos AI Scientist platform (Edison Scientific; access dates February–April 2026) to execute pre-specified bulk RNA-seq analytical workflows, including DESeq2 differential expression, fgsea enrichment and correlation analysis. All analytical pipelines, parameters, gene-set definitions, and statistical thresholds were specified by the authors prior to execution; all Kosmos outputs were independently reviewed, validated against source data, and interpreted by the authors. During manuscript preparation, the authors used ChatGPT (OpenAI; access dates [April 2026] for limited language editing; ChatGPT was not used to generate data, perform analyses, or inform scientific conclusions. All scientific content, interpretations, and conclusions were developed by the authors, who take full responsibility for the integrity of the work.

